# When can predictive uncertainty be trusted? A methodological evaluation in free-living wearable electrocardiogram signal-quality assessment

**DOI:** 10.64898/2026.08.25.26361304

**Authors:** Khanh Duy Tran

## Abstract

Uncertainty quantification is proposed as a safeguard for machine-learning systems in health-related signal analysis, but an uncertainty score is useful only if it behaves as a reliability signal. Free-living wearable electrocardiogram (ECG) signal-quality assessment provides a test bed because ambiguity, artifact, and acquisition shift can alter the relationship between confidence and correctness. This study evaluates predictive uncertainty under ambiguity, controlled corruption, and external distribution shift. 32,224 non-overlapping 10-s windows of synchronised single-lead ECG and three-axis accelerometry from 15 subjects in the Brno University of Technology ECG Quality Database were analysed. Two model families were compared: multinomial logistic regression and Classification and Regression Tree (CART), each progressing from a point estimate to a fixed-structure posterior and then a structure posterior. Expected conditional entropy and mutual information were evaluated as designated aleatoric and epistemic uncertainty measures, with max-softmax uncertainty as a confidence baseline. Validation covered error ranking, selective prediction, behavioural probes, posterior structural diversity, recorded-noise stress testing, and zero-shot external transfer. The logistic structure posterior retained an expected 8.5 of nine features and concentrated on near-complete masks, yielding little additional predictive diversity. Bayesian CART produced 221 distinct complete topologies among 238 retained draws and stronger score-dependent selective-risk behaviour. Conditional entropy increased with local class overlap, whereas mutual information increased when training information was reduced, although both showed cross-sensitivity. Under recorded noise, predicted quality severity changed more consistently than uncertainty, while external transfer preserved ordinal severity more reliably than uncertainty ordering. These findings show that posterior richness alone does not establish reliable uncertainty. Model-derived uncertainty should therefore be validated against prespecified ambiguity, information, and shift probes before supporting abstention, reacquisition, or downstream decisions.

## 1 Introduction

Uncertainty quantification (UQ) is increasingly used to support reliability assessment in medical artificial intelligence and physiological-signal analysis, particularly when a model may encounter ambiguous observations or data that differ from its development distribution (Begoli et al., 2019; Kompa et al., 2021; Ovadia et al., 2019). However, the existence of a posterior distribution or an uncertainty score does not by itself demonstrate that uncertainty is useful. For an uncertainty estimate to support abstention, reacquisition or human review, it should show an empirically defensible relationship with prediction error, observation ambiguity, limited training information or distribution shift. This makes uncertainty quantification not only a modelling problem but also a methodological validation problem.

Wearable electrocardiogram (ECG) signal-quality assessment provides a suitable test bed for this question. Ambulatory recordings are affected by electrode contact, motion, posture, muscle activity, power-line interference and subject-specific morphology, while a classifier may still return a high-probability label for an unreliable or unfamiliar segment. The Brno University of Technology ECG Quality Database (BUT QDB) contains free-living wearable ECG with synchronised accelerometry and three expert-defined quality levels, ranging from interpretable morphology to recordings in which reliable QRS detection is no longer possible (Nemcova et al., 2020). Previous work on BUT QDB has primarily established that signal quality can be discriminated using hand-crafted classifiers, deep sequence models and anomaly-detection approaches (Ma et al., 2022; Liu et al., 2022; Ma et al., 2023; Dua et al., 2022; Fan et al., 2026; Ibrahim et al., 2026). Those studies address whether quality can be predicted; they do not establish whether the uncertainty attached to an individual prediction remains meaningful when observations are ambiguous, corrupted or shifted.

This distinction exposes a methodological gap. First, it is unclear whether posterior-derived uncertainty provides useful failure ranking beyond the much simpler uncertainty implied by the maximum predicted class probability. Second, common categorical decompositions into expected conditional entropy and mutual information are frequently interpreted as aleatoric and epistemic uncertainty, yet their empirical separation is not guaranteed because both may respond to class overlap, label ambiguity and the amount of training information (Kendall and Gal, 2017; Depeweg et al., 2018; Hüllermeier and Waegeman, 2021; Wimmer et al., 2023). Third, uncertainty that behaves plausibly in-distribution may not preserve that behaviour under realistic artifact or external acquisition shift (Ovadia et al., 2019). Consequently, a useful evaluation should test uncertainty against explicit behavioural probes rather than infer reliability from model class or Bayesian formulation alone.

Several complementary criteria are available for such validation. Calibration error measures the agreement between confidence and empirical correctness (Guo et al., 2017), while negative log-likelihood and the Brier score are proper scoring rules for the complete predictive distribution (Gneiting and Raftery, 2007). Maximum softmax probability provides a conventional baseline for misclassification and out-of-distribution detection (Hendrycks and Gimpel, 2017). Selective classification additionally tests whether uncertainty can order cases so that risk decreases when the most uncertain predictions are rejected (Geifman and El-Yaniv, 2017; Traub et al., 2024). Observation ambiguity can be probed separately through expert disagreement and local class overlap, which retain information lost when disagreement is collapsed to a consensus label (Uma et al., 2021; Baan et al., 2022; Hüllermeier and Waegeman, 2021; Gruber et al., 2023). Together, these criteria allow uncertainty to be evaluated as an empirical construct rather than a descriptive property of a fitted model.

To keep this validation interpretable, the input representation is deliberately compact and auditable. Established ECG signal-quality indices describe beat detectability, morphology, spectral contamination and signal complexity (Clifford et al., 2012; Orphanidou et al., 2015; Zhao and Zhang, 2018), while synchronised accelerometry has been used to characterise wearable motion and its relationship with ECG degradation (Hamidi et al., 2023; Beach et al., 2021; Lindsey et al., 2025). Features motivated by these mechanisms are grouped, screened with a point-estimate model and then locked before uncertainty modelling so that the subsequent experiments primarily interrogate the uncertainty models rather than changing the representation.

The uncertainty models are organised as matched three-rung logistic and tree families. The logistic family progresses from maximum-likelihood coefficients (A0), to a fixed-feature Laplace coefficient posterior (A1) (Tierney and Kadane, 1986), and then to a spike-and-slab posterior over active-feature structures and coefficients (A2) (George and McCulloch, 1993). The Classification and Regression Tree (CART) family progresses from empirical leaf probabilities on one fitted partition (B0), to Dirichlet leaf-probability posteriors with that partition fixed (B1), and finally to posterior averaging over tree structures and leaves (B2) (Chipman et al., 1998). This matched progression creates a controlled comparison between uncertainty within a fixed structure and uncertainty over model structure, allowing posterior richness to be related directly to predictive diversity and operational reliability.

Accordingly, the objective of this study is not to demonstrate that Bayesian models are uniformly more accurate, nor to claim direct clinical benefit. The objective is to determine under what conditions model-derived uncertainty can be defended as a reliability signal for wearable ECG signal-quality assessment. Four prespecified questions are examined:

1. **Incremental reliability:** Does posterior-derived uncertainty improve error ranking and selective rejection beyond same-model max-softmax uncertainty?
2. **Construct behaviour:** Do the designated aleatoric and epistemic components respond in the expected directions to observation ambiguity and reduced training information, and where do they show cross-sensitivity?
3. **Posterior structure:** Does posterior support concentrate on a stable representation or remain structurally diffuse, and does that structural diversity translate into predictive disagreement?
4. **Robustness under shift:** Does in-distribution uncertainty behaviour persist under recorded ECG corruption and zero-shot transfer to the Long-Term ST Database (LT-STDB)?

By treating ECG quality classification as a controlled application rather than the endpoint of the study, the analysis tests when uncertainty quantification provides additional reliability information and when it does not.

## 2 Materials and methods

Figure 1 summarises the two-stage design: a common ECG—accelerometer representation is locked first, then matched logistic and tree ladders progress from point estimation to fixed-structure and structure posteriors before reliability is evaluated in distribution and under shift.

**Figure 1.**
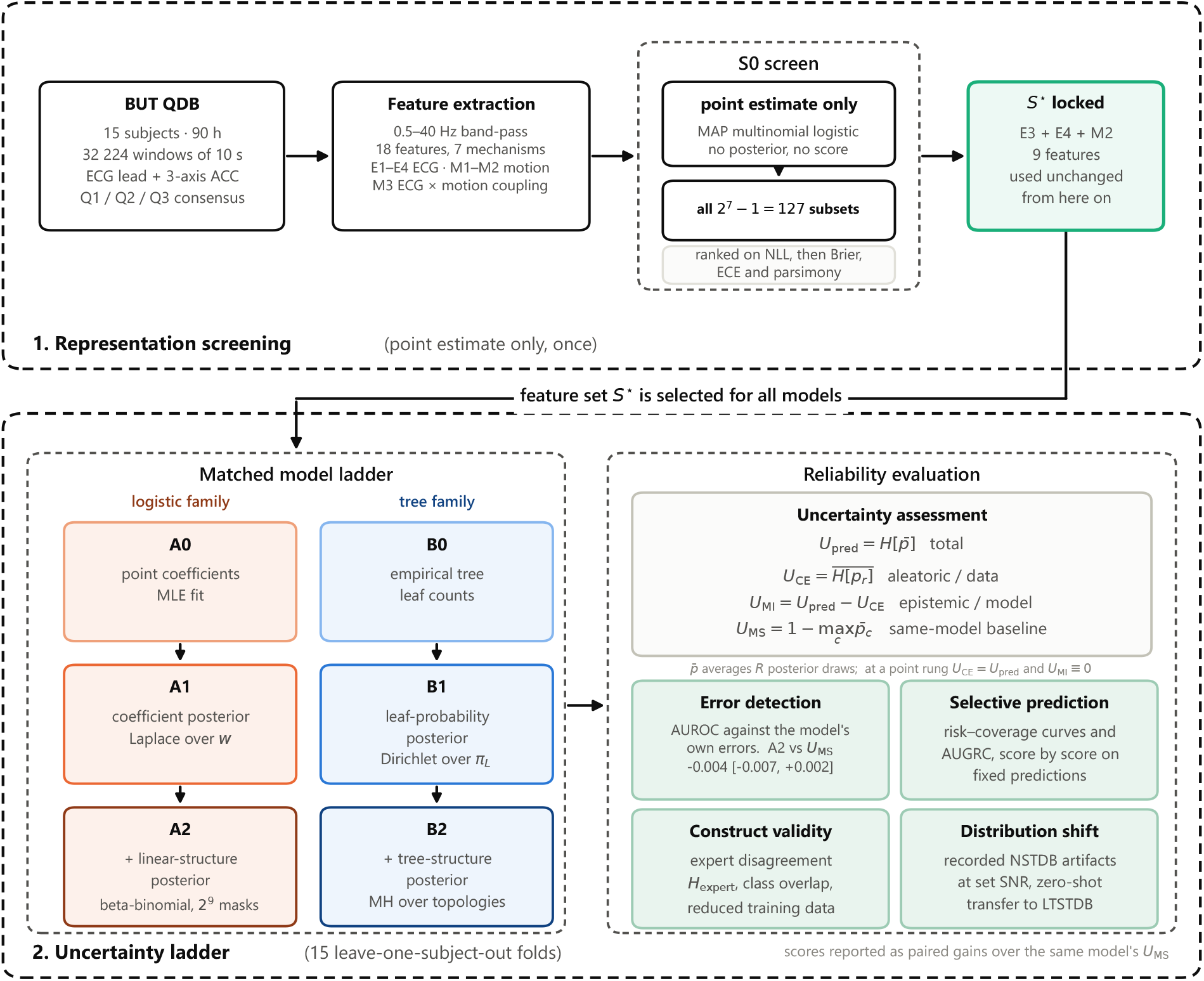
Two-stage study design. Stage 1 screens seven ECG/accelerometer mechanism groups and locks *S*^⋆^; Stage 2 compares matched logistic (A0—A2) and tree (B0—B2) ladders from point estimation to fixed-structure and structure posteriors, followed by reliability and shift evaluation.

### 2.1 Databases and sensing modalities

BUT QDB is the development and leave-one-subject-out (LOSO) evaluation cohort. The Massachusetts Institute of Technology—Beth Israel Hospital (MIT-BIH) Noise Stress Test Database (NSTDB) supplies recorded ECG artifacts for the controlled stress test, and LTSTDB is used only for frozen zero-shot transfer. Neither NSTDB nor LTSTDB contributes fitting data or matched three-class labels for accuracy evaluation.

#### BUT QDB

The Brno University of Technology ECG Quality Database contains 18 free-living recordings from 15 subjects acquired with a Bittium Faros 180 mobile recorder, providing synchronised single-lead ECG (1000 Hz) and integrated three-axis accelerometry (100 Hz), with each recording lasting at least 24 h (Nemcova et al., 2020; Goldberger et al., 2000). This synchronised motion channel is unusual in ECG-quality databases and permits explicit testing of motion-related features (van der Bijl et al., 2022). Three experts independently annotated quality; their consensus is the classification target and the individual votes are retained only for the disagreement analysis. Quality has three ordered classes: Q1, morphology interpretable; Q2, morphology degraded but QRS detection reliable; and Q3, reliable QRS detection no longer possible.

#### NSTDB

The MIT-BIH Noise Stress Test Database supplies independently recorded baseline-wander, electrode-motion and muscle-artifact traces for the controlled stress test. The artifact recordings were acquired from physically active volunteers using a Holter ECG recorder, standard leads and electrodes, with two channels recorded simultaneously (Moody et al., 1984; Goldberger et al., 2000). NSTDB is used as a noise library, not as labelled ECG; the injection protocol is defined in Section 2.7.

#### LTSTDB

The Long-Term ST Database (LTSTDB) contains 86 long-term ambulatory recordings from 80 subjects, 1,992 h in total, with two or three ECG channels digitised at 250 Hz and no synchronised accelerometry (Jager et al., 2003; Goldberger et al., 2000). The source recordings were collected in routine clinical settings across multiple sites using standard ambulatory ECG recorders and several digitisation systems: 68 records are two-channel and 18 are three-channel, and each runs for roughly 19—26 h (Jager et al., 2003). Zero-shot transfer therefore uses 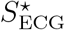 on the first recorded channel of each record.

That channel is the unit LTSTDB’s own annotations address — the database numbers the channels of a record and writes quality marks against those numbers, so an unreadable interval on the first channel is recorded as `urd0` (Jager et al., 2003). Fixing the first channel is therefore a rule stated in the database’s own terms, and it was applied before any transfer result was inspected, but it is not a fixed physical lead. Across the 86 records the first channel is a precordial lead in 26 (V4 in 21, V5 in 2, V6 in 2, V2 in 1), a bipolar limb-type lead in 23 (modified limb lead L2 (ML2) in 16 and modified limb lead III (MLIII) in 7), an orthogonal E-S lead in 15, and unnamed in the remaining 22. Among the seven records that carry a bounded unreadable interval it is V4 in three, E-S in two, and MLIII and V5 in one each.

Electrode placement is thus heterogeneous within the transfer target as well as different from BUT QDB’s single chest lead, which is part of what makes this a composite rather than a controlled shift.

### 2.2 Windows, labels and validation protocol

The ECG is decimated to 250 Hz (Nyquist 125 Hz, which retains both the mains and electromyographic bands) and the accelerometer is kept at its native 100 Hz. Signals are divided into non-overlapping 10-s windows. A window enters the study only when the consensus annotation covers all of it and at least 90% of its samples carry the modal class; that modal class is the label. This yields 32,224 windows, 89.5 h, from 15 subjects (Table 1). The class distribution is 16,687 Q1, 10,292 Q2 and 5,245 Q3. The cohort is severely unbalanced at the subject level: subjects 105, 100 and 111 contribute 29,194 of the 32,224 windows, and subject 105 alone contributes 4,734 of the 5,245 Q3 windows.

**Table 1.**
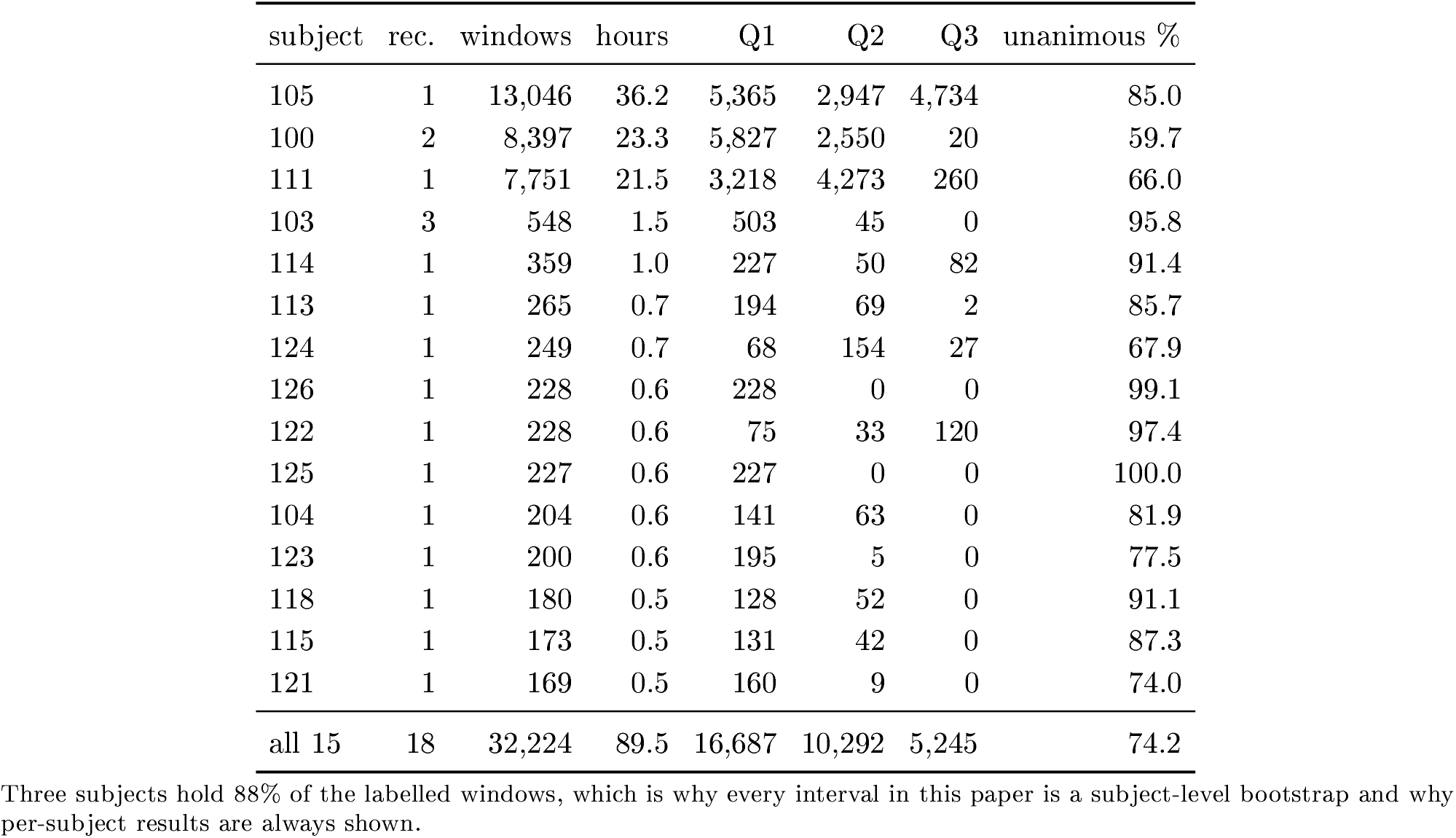
BUT QDB windows entering the study, by subject. A window is used when the consensus annotation covers all of it and at least 90% of its samples carry the modal class.

| subject | rec. | windows | hours | Q1 | Q2 | Q3 | unanimous % |
| --- | --- | --- | --- | --- | --- | --- | --- |
| 105 | 1 | 13,046 | 36.2 | 5,365 | 2,947 | 4,734 | 85.0 |
| 100 | 2 | 8,397 | 23.3 | 5,827 | 2,550 | 20 | 59.7 |
| 111 | 1 | 7,751 | 21.5 | 3,218 | 4,273 | 260 | 66.0 |
| 103 | 3 | 548 | 1.5 | 503 | 45 | 0 | 95.8 |
| 114 | 1 | 359 | 1.0 | 227 | 50 | 82 | 91.4 |
| 113 | 1 | 265 | 0.7 | 194 | 69 | 2 | 85.7 |
| 124 | 1 | 249 | 0.7 | 68 | 154 | 27 | 67.9 |
| 126 | 1 | 228 | 0.6 | 228 | 0 | 0 | 99.1 |
| 122 | 1 | 228 | 0.6 | 75 | 33 | 120 | 97.4 |
| 125 | 1 | 227 | 0.6 | 227 | 0 | 0 | 100.0 |
| 104 | 1 | 204 | 0.6 | 141 | 63 | 0 | 81.9 |
| 123 | 1 | 200 | 0.6 | 195 | 5 | 0 | 77.5 |
| 118 | 1 | 180 | 0.5 | 128 | 52 | 0 | 91.1 |
| 115 | 1 | 173 | 0.5 | 131 | 42 | 0 | 87.3 |
| 121 | 1 | 169 | 0.5 | 160 | 9 | 0 | 74.0 |
| all 15 | 18 | 32,224 | 89.5 | 16,687 | 10,292 | 5,245 | 74.2 |

Three subjects hold 88% of the labelled windows, which is why every interval in this paper is a subject-level bootstrap and why per-subject results are always shown.

Expert vote entropy *H*_expert_, defined in Section 2.6, is retained as an auxiliary measure of annotation ambiguity rather than as a model input. In multi-annotator datasets, majority voting is commonly used to obtain a single target label, but this aggregation can discard information contained in disagreement among annotators (Uma et al., 2021; Baan et al., 2022). Accordingly, the distribution of individual expert ratings can provide an additional indication of instance-level ambiguity, with entropy offering a natural summary of the dispersion of those ratings (Baan et al., 2022). A window is termed *unanimous* when all three experts assign the same quality label, and *non-unanimous* when at least one expert assigns a different label. Of the 32,224 windows, 8,320 (25.8%) are non-unanimous. Because both the consensus target label and *H*_expert_ are derived from the same expert ratings, *H*_expert_ is independent of the fitted model but is not statistically independent of target construction.

Validation uses leave-one-subject-out (LOSO) cross-validation over 15 subjects, with standardisation estimated from each training fold only. Confidence intervals use 1000 paired subject-level bootstrap resamples rather than window resampling because the cohort is strongly imbalanced by subject.

### 2.3 Representation screening

The candidate feature universe was chosen for simplicity, interpretability and prior use in ECG signal-quality assessment. Representation screening selects among these predefined mechanism groups with a point-estimate model; the selected columns are then held fixed as the common input space for the matched uncertainty experiment.

#### Candidate mechanisms

Eighteen interpretable scalars are grouped into seven predefined mechanisms: ECG detectability/morphology (E1), rhythm plausibility (E2), spectral contamination (E3), statistical complexity (E4), accelerometer motion magnitude (M1), motion dynamics (M2), and cross-modal ECG—motion coupling (M3). The groups are based on established ECG signal-quality, accelerometry and electrode-motion work (Clifford et al., 2012; Orphanidou et al., 2015; Zhao and Zhang, 2018; Zhang et al., 2015; van Hees et al., 2013; Vähä-Ypyä et al., 2015; Bakrania et al., 2016; Kojima et al., 2008; Beach et al., 2021). M3 requires both signals and adapts the velocity-related coupling strategy of Beach et al. (2021) to 10-s BUT QDB windows. The definitions and implementation conventions for the selected representation are given explicitly below.

#### Mechanism-subset screening

All 2^7^ − 1 = 127 non-empty mechanism subsets were evaluated with S0, a maximum a posteriori (MAP) multinomial-logistic point model, using a five-fold subject-grouped development split; its penalty was selected by inner grouped crossvalidation. Screening was performed at the mechanism level without using uncertainty end-points. Subsets were ranked by negative log-likelihood (NLL), then Brier score, then expected calibration error (ECE), with parsimony as the final tie-break. The selected representation was then fixed before the uncertainty model comparison.

#### Selected representation

The screen selected *S*^⋆^ = E3 + E4 + M2, comprising nine features spanning ECG spectral contamination, ECG statistical complexity, and accelerometer motion dynamics. E3 contains the power spectral quality index (pSQI), baseline-wander spectral ratio (basSQI), high-frequency contamination ratio *r*_HF_, and 50-Hz mains ratio *r*_50_; E4 contains the kurtosis signal-quality index (kSQI) and multiscale sample entropy (MSE); and M2 contains accelerometer-magnitude jerk, dominant frequency *f*_dom_, and normalised spectral entropy *H*_spec_. The locked input vector is

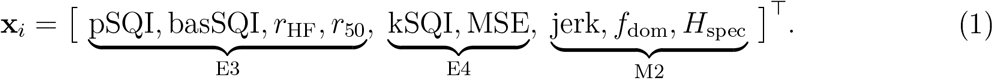

Because these nine features constitute the fixed representation used by all subsequent uncertainty models, their definitions and implementation conventions are given explicitly below. The ECG spectral-quality descriptors in E3 and the statistical-quality descriptors in E4 follow established ECG signal-quality formulations (Clifford et al., 2012; Orphanidou et al., 2015; Zhao and Zhang, 2018), with multiscale entropy used to characterise signal complexity (Zhang et al., 2015). The M2 descriptors characterise wearable motion dynamics using accelerometer magnitude, temporal variation, and spectral concentration, consistent with prior accelerometry and wearable-artifact studies (Kojima et al., 2008; Hamidi et al., 2023; Beach et al., 2021; Lindsey et al., 2025).

Let 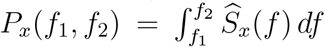 denote ECG Welch band power, 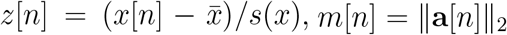, and 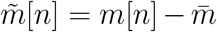. Here SampEn(·) denotes sample entropy. The selected features are

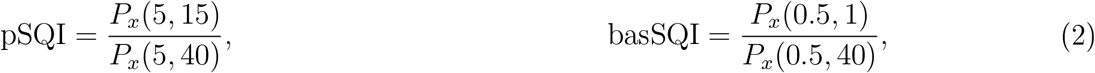

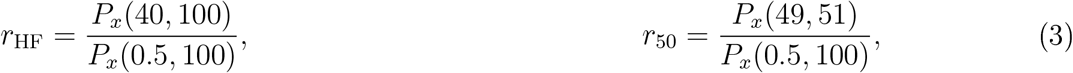

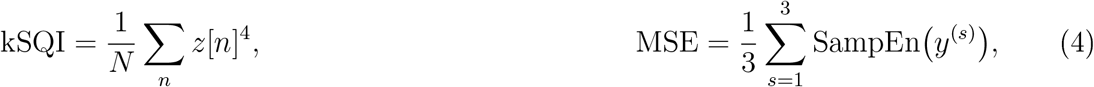

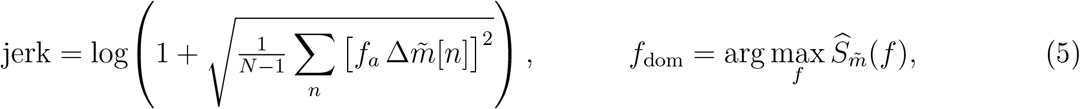

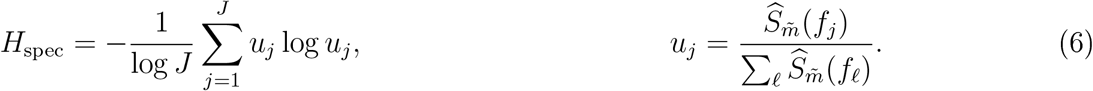

Here *y*^(*s*)^ denotes the ECG coarse-grained at scale *s* after decimation to 50 Hz, sample entropy is evaluated with embedding dimension *m* = 2 and tolerance *r* = 0.15 *s*(*y*), 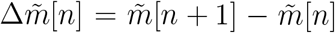, *f*_*a*_ = 100 Hz is the accelerometer sampling rate, and 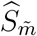 is the Welch spectrum of the mean-removed accelerometer magnitude. The ECG-only subset, 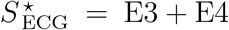 contains six features and is used for the ECG-only stress-test arm and LTSTDB transfer, where accelerometer measurements are unavailable.

All six rungs use this same nine-feature universe. A0, A1, B0 and B1 use *S*^⋆^ directly; A2 averages over subsets of these nine admitted features, while B2 averages over tree partitions of the same columns. Because representation screening and the leave-one-subject-out experiment use the same 15-subject cohort, absolute performance conditional on *S*^⋆^ remains a post-selection estimate. The paired rung-to-rung contrasts are therefore the primary inferential target.

### 2.4 Matched probabilistic model ladder

The two families are matched by the object over which uncertainty is represented. Maximum-likelihood estimation (MLE) is used to obtain the coefficient point estimate for A0.

| stage | logistic family | tree family |
| --- | --- | --- |
| point predictor | A0: MLE coefficients | B0: empirical CART leaves |
| fixed-structure posterior | A1: coefficient posterior | B1: leaf-probability posterior |
| structure posterior | A2: feature-mask + coefficients | B2: topology + leaf probabilities |

The progression is point estimate → posterior within a fixed structure → posterior over structure. This is the same general motivation that underlies multi-predictor uncertainty methods such as Monte Carlo dropout and deep ensembles, although those methods generate predictive diversity differently (Gal and Ghahramani, 2016; Lakshminarayanan et al., 2017). Here the posterior construction is explicit: A1/B1 quantify uncertainty conditional on one structure, whereas A2/B2 average predictions over alternative structures as well (Wilson and Izmailov, 2020; Chipman et al., 1998).

#### Logistic family

All three rungs use the same multinomial softmax likelihood. For augmented feature vector 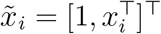 and class parameters *θ*_*c*_,

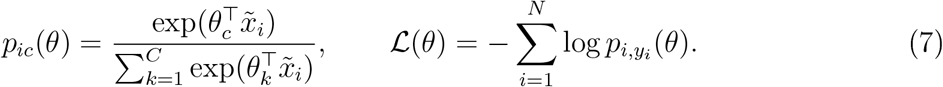

A0 is the literal maximum-likelihood fit, 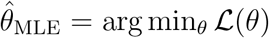, with no prior term at all. Its predictive distribution is therefore the single fitted softmax

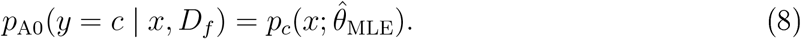

A near-unpenalised ridge would be a maximum-a-posteriori fit under a diffuse prior, and the A1–A0 contrast would then compare two prior strengths rather than test whether a prior is present; the optimiser’s behaviour at *α*_0_ = 0 is measured and reported in Section 3.2 rather than avoided.

A1 places an isotropic Gaussian prior *θ* ~ *N* (0, *α*^−1^*I*) on the coefficients. The slab precision is selected by empirical Bayes within each training fold, 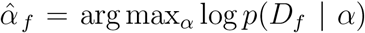, using the same Laplace approximation as the posterior. The fold-specific 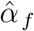 is reused unchanged in A2, so A1—A2 isolates the addition of feature-structure uncertainty rather than a change in slab precision. A1 represents that posterior by a Laplace approximation about the mode (Tierney and Kadane, 1986),

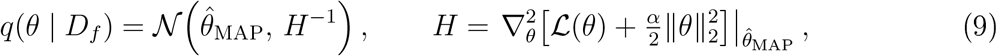

from which *R* = 100 parameter vectors are drawn. The corresponding posterior predictive distribution is

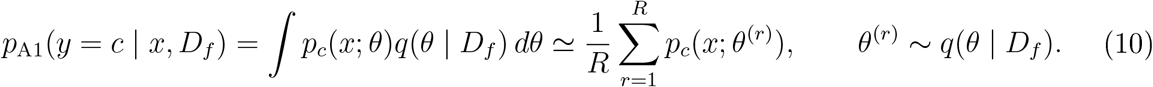

The Hessian is formed exactly; the Gaussian posterior shape is the approximation.

A2 is a spike-and-slab logistic model: each of the nine features of *S*^⋆^ carries an inclusion indicator, and the posterior is joint over which features are active and what their coefficients are,

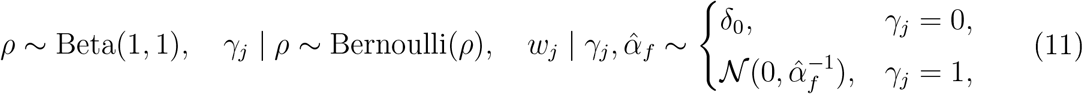

with *ρ* integrated analytically, giving a beta-binomial inclusion prior that places equal mass on each model *size* rather than on each model. That multiplicity correction matters: there are 126 masks of size four and one of size nine, so a flat prior over masks would concentrate before any data were seen. The predictive distribution is

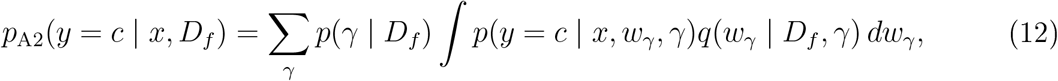

where *q*(*w*_*γ*_ | *D*_*f*_, *γ*) is the structure-specific Laplace posterior. Thus A2 averages both coefficient uncertainty and uncertainty over the active-feature structure.

One indicator governs each feature across all classes, so *γ*_*j*_ = 0 removes feature *j* from the multinomial model entirely rather than from one class’s logit; a structure is therefore a statement about which measurements the model uses. With nine features the space has 2^9^ = 512 members including the intercept-only structure, which is small enough to enumerate, so the posterior over *γ* is computed exactly rather than explored by Markov chain Monte Carlo (MCMC). Each structure is weighted by its Laplace-approximated marginal likelihood. Up to constants common to all structures,

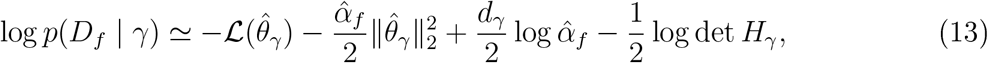

where 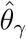 is the structure-specific MAP mode, *d*_*γ*_ is the number of coefficients governed by the Gaussian slab, and *H*_*γ*_ is the corresponding negative-log-posterior Hessian. The last two terms provide the Laplace complexity correction, preventing larger structures from being favoured solely because they contain more parameters. A1 is the *γ* = **1** special case of A2.

The structure space is closed under *S*^⋆^: A2 chooses which admitted features the linear model needs and cannot readmit a mechanism excluded by the screening step. The screening step fixes the information available; the structure posterior decides only how that information is organised.

#### Tree family

Let *T* denote a tree, *L*(*x*; *T*) the terminal leaf reached by *x, n*_*Lc*_ the number of training observations of class *c* in leaf *L*, and *n*_*L*_ = ∑_*c*_ *n*_*Lc*_. B0 is a Classification and Regression Tree (CART) classifier with maximum depth 12 and minimum leaf size 10. Its predictive probabilities are the empirical leaf frequencies,

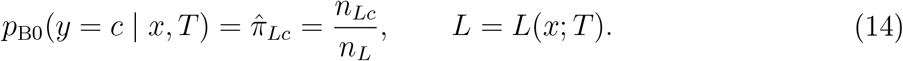

Classes absent from a training leaf receive *ε* = 10^−12^ before renormalisation only when a finite log score is required; the realised class probability is clipped to [*ε*, 1]. No probability smoothing is otherwise applied, so B0 remains the empirical point-estimate control.

B1 holds the B0 topology fixed and replaces each empirical leaf distribution by a Dirichlet— multinomial posterior,

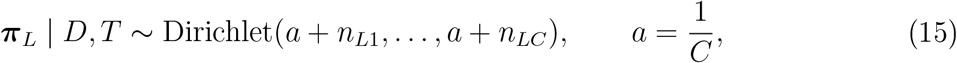

with *R* = 100 posterior leaf-probability draws used for prediction. Conditional on the fixed tree, the posterior predictive mean is

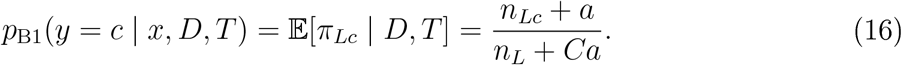

B1—B0 therefore isolates uncertainty and smoothing in the leaf probabilities while keeping the partition unchanged.

B2 additionally averages over the partition itself. Its posterior predictive distribution is

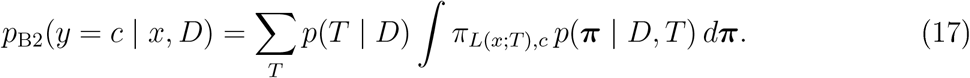

Following Bayesian CART (Chipman et al., 1998), the topology prior uses *p*(split at depth *d*) 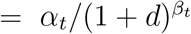 with *α*_*t*_ = 0.95 and *β*_*t*_ = 2, a uniform split prior over a per-feature quantile grid fixed before sampling, and the same Dirichlet leaves as B1. Grow, prune and change moves are accepted by Metropolis—Hastings. Eight independent chains of 2000 iterations are pooled after 500 burn-in iterations and thinning by 50. Thus *U*_MI_ at B2 measures posterior predictive disagreement induced jointly by uncertainty in tree structure and leaf probabilities, directly paralleling the feature-structure and coefficient uncertainty averaged in A2.

### 2.5 Predictive uncertainty measures

For *R* posterior predictive members *p*_*r*_(*c*) = *p*(*y* = *c* | *x, θ*_*r*_), define the uncertainty quantities below. The subscripts CE, MI and MS denote conditional entropy, mutual information and max-softmax, respectively.

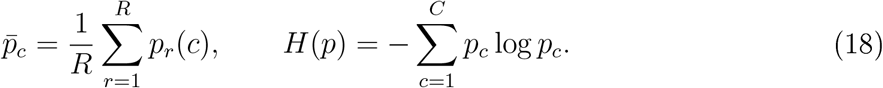

Predictive entropy is decomposed as

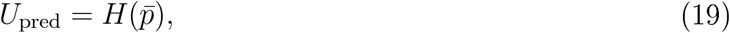

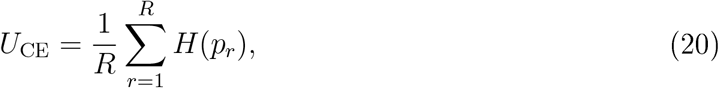

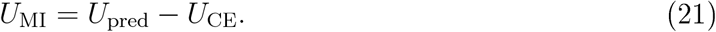

Throughout this study, *U*_CE_ is the *aleatoric/data uncertainty component:* it is the posterior expectation of within-model class ambiguity for the observed feature vector. *U*_MI_ is the *epistemic/model uncertainty component:* it is the mutual information between the predicted class and the posterior model draw, and therefore measures disagreement induced by uncertainty in coefficients, feature structure, leaf probabilities or tree structure, depending on the rung. At a point-estimate rung (*R* = 1), *U*_CE_ = *U*_pred_ and *U*_MI_ ≡ 0 by construction.

These labels follow the conventional predictive-entropy decomposition (Depeweg et al., 2018; Kendall and Gal, 2017; Hüllermeier and Waegeman, 2021): predictive entropy measures total class ambiguity, expected conditional entropy averages within-member ambiguity, and mutual information measures disagreement across posterior members. The same quantities are widely used in Bayesian and biosignal uncertainty quantification (UQ) (Vranken et al., 2021; de Jong et al., 2026; Bench et al., 2026).

The components are used operationally rather than assumed to be perfectly separable physical sources: their behaviour can depend on model class and posterior representation (Wimmer et al., 2023; Gruber et al., 2023). We therefore test the designated roles directly. An aleatoric reading predicts increasing *U*_CE_ with observation ambiguity; an epistemic reading predicts increasing *U*_MI_ when training information is reduced, while changes in the non-target component reveal cross-sensitivity.

Max-softmax uncertainty provides the same-model baseline (Hendrycks and Gimpel, 2017), oriented so that larger values mean less certainty:

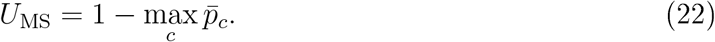

Posterior-derived scores are evaluated by whether they add reliability beyond this inexpensive confidence-based signal.

### 2.6 Reliability metrics and validation probes

Reliability is assessed on complementary axes rather than by a single score (Ovadia et al., 2019; de Jong et al., 2026; Bench et al., 2026). Predictive performance is summarised by macro-F1 and balanced accuracy; probability quality by NLL and multiclass Brier score; and calibration by 15-bin equal-width ECE (Gneiting and Raftery, 2007; Guo et al., 2017). Standard definitions are used for these metrics. The primary uncertainty endpoints are the area under the receiver operating characteristic curve for error detection (error-detection AUROC), which tests whether uncertainty ranks errors above correct predictions, and the area under the generalized risk—coverage curve (AUGRC), which measures the resulting undetected-failure burden under rejection (Hendrycks and Gimpel, 2017; Geifman and El-Yaniv, 2017; Traub et al., 2024). Expert and local-label entropy are used as ambiguity probes (Uma et al., 2021; Baan et al., 2022; Vranken et al., 2021; Hüllermeier and Waegeman, 2021; Wimmer et al., 2023).

For an uncertainty score *U*_*i*_ (larger means less certain), error-detection AUROC is the ranking probability

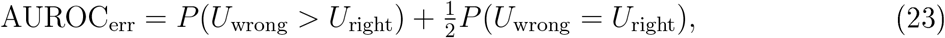

Annotator disagreement is represented by expert vote entropy. Let *y*_*ir*_ be the class assigned to window *i* by expert *r*, with *R* = 3 experts, and let

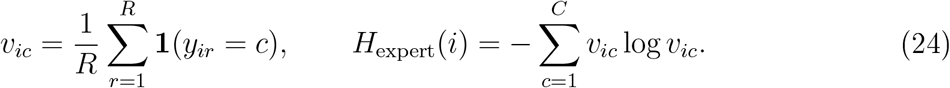

With three experts, *H*_expert_ is 0 for unanimity, 0.6365 nats for a two-to-one split and 1.0986 nats for three different votes. We report within-subject Spearman *ρ*_*s*_(*U, H*_expert_) and the mean uncertainty difference between non-unanimous and unanimous windows. Because the consensus label and *H*_expert_ use the same ratings, this is an ambiguity probe rather than an independent reference.

Selective prediction follows the noise-free selective-classification framework (El-Yaniv and Wiener, 2010; Geifman and El-Yaniv, 2017): sort windows so that *U*_(1)_ ≤ · · · ≤ *U*_(*N*)_. At accepted coverage *κ*_*k*_ = *k/N*,

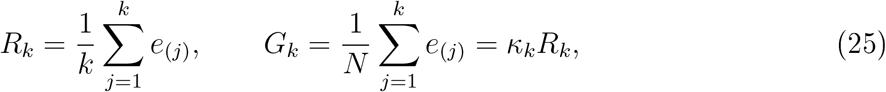

The primary area summary is

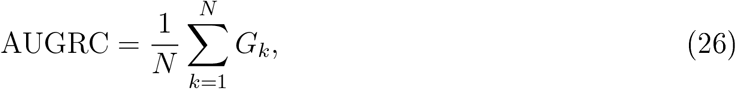

which averages the proportion of undetected failures across coverage levels (Geifman and El-Yaniv, 2017; Geifman et al., 2019; Traub et al., 2024).

All confidence intervals use 1000 paired subject-level bootstrap resamples; paired contrasts are recomputed within each resample.

#### Incremental reliability over *U*_MS_

The deployment question is whether a posterior-derived score carries information beyond the confidence already available from the same predictive probabilities. Using *U*_MS_ from Equation (22), for *U* ∈ {*U*_pred_, *U*_CE_, *U*_MI_} we define

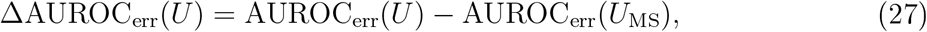

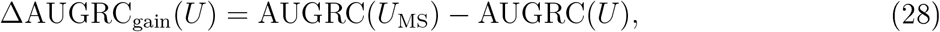

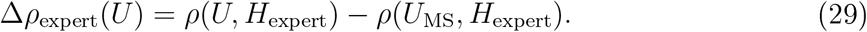

Signs are oriented so that positive values favour the posterior-derived score. Because each contrast uses identical model predictions, it isolates the uncertainty ranking rather than predictive accuracy.

#### Validation probes for aleatoric and epistemic roles

The signed expectations in Section 2.5 are tested with three prespecified probes. Expert vote entropy and local class overlap probe observation ambiguity for *U*_CE_. For each held-out window *x*_*i*_, the *k* nearest training windows in the locked standardised feature space estimate

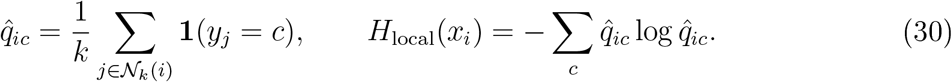

Larger *H*_local_ indicates stronger local label overlap. Neighbours are drawn only from the corresponding training fold, with *k* = 50 fixed throughout the analysis.

The epistemic probe refits each posterior rung on nested fractions of its training fold; *U*_MI_ is expected to rise as training information decreases (Wimmer et al., 2023; Gruber et al., 2023). Both components are reported for every probe to expose cross-sensitivity.

Test-time ECG corruption is not used for this construct test. Because the prediction target is ECG signal quality, contaminating a window can change its true quality class as well as the observation. The recorded-noise sweep is therefore treated separately as a distribution-shift experiment (Section 2.7).

### 2.7 Controlled ECG corruption and external transfer

#### Standardised ECG noise stress

Baseline wander, electrode motion and muscle artifact from NSTDB were resampled to 250 Hz and added separately to held-out BUT QDB ECG (Moody et al., 1984; Goldberger et al., 2000).

For a held-out BUT QDB ECG window *x*_*i*_[*n*] and an independently offset NSTDB artifact segment *n*_*i*_[*n*], both at 250 Hz, the corrupted signal at target signal-to-noise ratio (SNR) *s* is

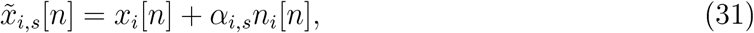

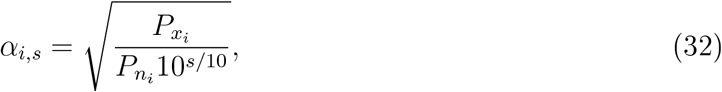

where 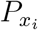 and 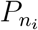 are the mean-centred window powers. Equivalently,

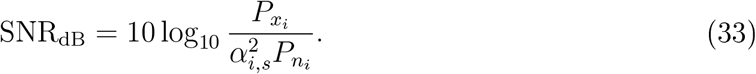

An independent random offset into the corresponding artifact trace is used for each window. The stress levels are clean (*α* = 0), 24, 18, 12, 6, 0 and –6 dB, and the three NSTDB artifact types are evaluated separately. Thus the noise severity is controlled while the underlying BUT QDB subject and recording context are held fixed.

Only ECG is perturbed; the synchronised accelerometer and LOSO assignment are unchanged. The test is run with *S*^⋆^ and with ECG-only *S*^⋆^ to check whether the untouched motion channel alters the response. No contaminated window enters fitting, representation selection or uncertainty-score selection; NSTDB is a shift test, not ground truth for aleatoric uncertainty.

Accuracy is not scored against the original BUT QDB label because added artifact can change the true signal-quality class. We therefore report predicted severity *S*_*Q*_ = 0 *p*(*Q*1) + 1 *p*(*Q*2)+2 *p*(*Q*3) and the uncertainty scores, separating response to degradation from response in uncertainty.

#### Zero-shot external transfer

LTSTDB does not provide Q1—Q3 labels, so its quality annotations are converted to study-defined window-level occupancy states. For a 10-s window *W*_*i*_ and the union U of bounded unreadable intervals,

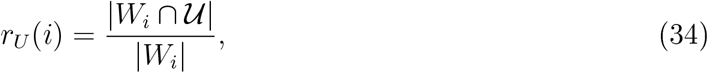

where *r*_*U*_ = 0 denotes clean, 0 *< r*_*U*_ *<* 1 transition, and *r*_*U*_ = 1 fully unreadable; transition is not treated as a surrogate for BUT Q2. Of the 86 LTSTDB recordings, 12 contain the quality annotations required for transfer and are segmented into non-overlapping 10-s windows at the native 250 Hz sampling rate. The first recorded channel contains 34 bounded unreadable intervals (18.34 h) across seven subjects. Because isolated noisy episodes are point-marked rather than bracketed (Jager et al., 2003), clean windows are additionally required to lie at least 30 s from any quality event, removing 288 windows. The resulting transfer set contains 99,110 scored windows (275.3 h): 92,480 clean, 63 transition, and 6,567 fully unreadable; 14 windows centred on isolated noisy instants are excluded from the transfer endpoints. Bounded unreadable intervals occur in seven subjects and are strongly concentrated in three, so inference is performed at the subject level rather than by pooling windows. The state-level summaries in Figure 9a,c require at least four unreadable windows and therefore retain all seven eligible subjects. Paired contrasts in Figure 9b use stricter support requirements: at least 20 windows for clean and unreadable states and at least four for transition, which is intrinsically sparse because it occurs only at interval boundaries. Consequently, transition-minus-clean and unreadable-minus-transition use *n* = 5, whereas unreadable-minus-clean uses *n* = 6.

All six rungs are then trained on all BUT QDB using 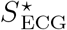, frozen, and applied to the first recorded LTSTDB channel with BUT-derived standardisation unchanged. No LT-STDB data enter fitting, calibration or threshold selection. The frozen models still output *p*(*Q*1), *p*(*Q*2), *p*(*Q*3), summarised by

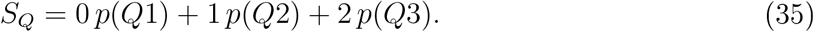

Transfer is tested by the prespecified ordering *S*_*Q*_(clean) *< S*_*Q*_(transition) *< S*_*Q*_(unreadable) and by the corresponding uncertainty scores; cross-database Q1—Q3 accuracy is not computed.

### 2.8 Implementation and reproducibility

All analyses were implemented in Python 3.13.5 using `NunPy` 2.3.2, `scikit-learn` 1.7.1, `pandas` 2.3.1 and `Hatplotlib` 3.10.5. Feature extraction, screening, model fitting, posterior inference, robustness analyses and figure generation were integrated into a single reproducible pipeline operating from the raw recordings. The logistic, Laplace, spike-and-slab and Bayesian CART procedures followed the formulations in Section 2.4.

Deterministic estimators were reproduced directly, whereas posterior-predictive sampling and Bayesian CART inference used fixed random seeds. Numerical checks were performed within each fold to verify optimisation, posterior approximation and sampling behaviour; the corresponding summaries are reported with the main results in Section 3.1.

## 3 Results

### 3.1 Representation screening and selected features

#### Mechanism-subset screen

Figure 2 summarises the 127-subset screen. Spectral contamination (E3) and motion dynamics (M2) appear in every one of the strongest configurations, statistical complexity (E4) in 62% of them, and motion magnitude (M1) in only 15%, against a size-matched null prevalence of 0.56. The selected representation *S*^⋆^ = E3 + E4 + M2 reaches NLL 0.414, Brier 0.235 and ECE 0.041 with nine features on the development split; the best eleven-feature configuration, which adds the cross-modal group M3, is worse on all three (0.420, 0.237, 0.043). Two results are worth stating plainly. Motion *dynamics* survives while motion *magnitude* does not, indicating that the temporal character of movement carries more useful information here than its overall amplitude. M3 was included on the basis of prior electrode-motion work, but adding its cross-modal coupling summaries to E3+E4+M2 does not improve held-out likelihood, Brier score or calibration in this cohort. Its exclusion is therefore interpreted as conditional redundancy within the selected representation, not as evidence that ECG—motion coupling lacks physiological or engineering relevance.

**Figure 2.**
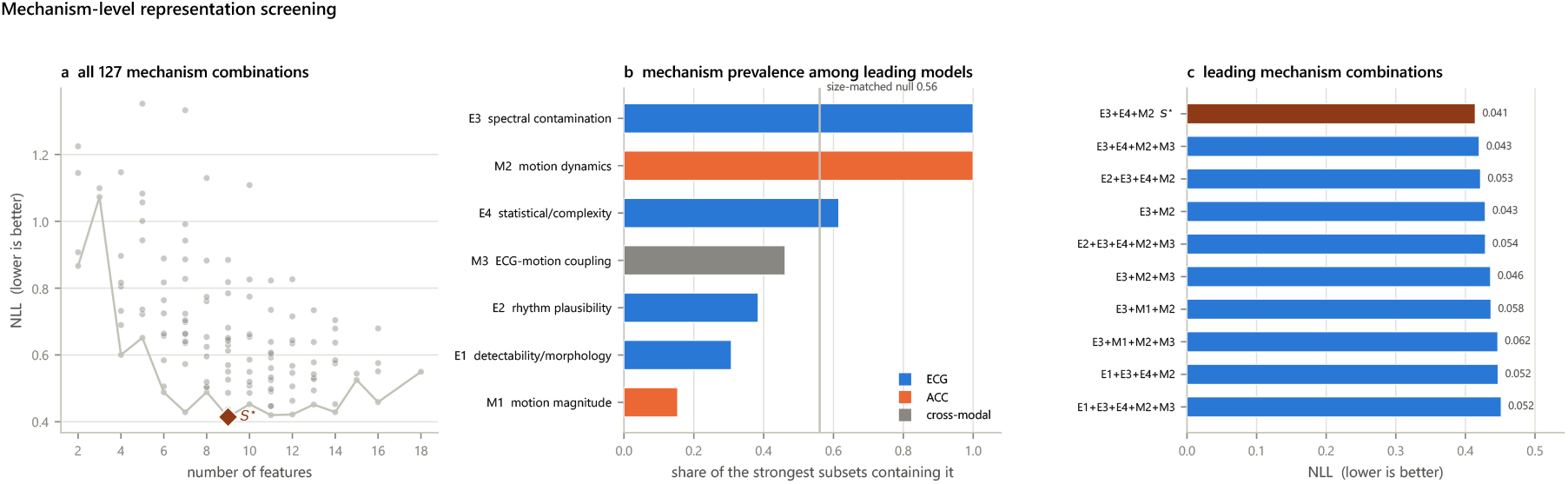
Mechanism-subset screen across all 127 non-empty mechanism subsets. Panels show subset NLL, mechanism prevalence among strong configurations and the leading configurations. The selected representation is *S*^⋆^ = E3 + E4 + M2.

The representation was therefore locked before any uncertainty endpoint was evaluated.

### 3.2 Predictive performance and posterior structure

#### Predictive performance

Table 2 and Figure 3 report the six rungs on *S*^⋆^, pooled over the out-of-subject predictions of all 15 folds. Absolute values carry selection optimism and should not be read as what a locked pipeline would achieve on a new cohort; the paired contrasts are the inferential target.

**Table 2.** LOSO predictive performance under the locked representation *S*^⋆^. Brackets are 95% subject-level bootstrap intervals. All six models receive the same locked nine-feature information universe; A2 and B2 additionally marginalise over model structure within that universe. Per-class recalls are in Supplementary Table S3.

| rung | model | macro-F1 | bal. acc. | NLL | Brier | ECE |
| --- | --- | --- | --- | --- | --- | --- |
| A0 | MLE logistic | 0.859<br>[0.541, 0.891] | 0.846<br>[0.541, 0.879] | 0.401<br>[0.227, 0.659] | 0.224<br>[0.136, 0.357] | 0.040<br>[0.013, 0.114] |
| A1 | Laplace logistic | 0.859<br>[0.541, 0.891] | 0.846<br>[0.541, 0.879] | 0.395<br>[0.223, 0.635] | 0.223<br>[0.134, 0.347] | 0.038<br>[0.013, 0.106] |
| A2 | spike-and-slab logistic | 0.860<br>[0.546, 0.892] | 0.846<br>[0.544, 0.879] | 0.394<br>[0.222, 0.634] | 0.222<br>[0.133, 0.347] | 0.038<br>[0.013, 0.106] |
| B0 | empirical CART | 0.815<br>[0.504, 0.865] | 0.810<br>[0.512, 0.859] | 2.045<br>[1.540, 3.217] | 0.303<br>[0.205, 0.429] | 0.114<br>[0.071, 0.196] |
| B1 | Dirichlet-leaf CART | 0.814<br>[0.501, 0.865] | 0.810<br>[0.510, 0.860] | 0.640<br>[0.423, 0.927] | 0.297<br>[0.201, 0.420] | 0.103<br>[0.063, 0.182] |
| B2 | Bayesian CART | 0.830<br>[0.501, 0.872] | 0.816<br>[0.507, 0.863] | 0.546<br>[0.469, 0.627] | 0.316<br>[0.252, 0.371] | 0.114<br>[0.041, 0.193] |

**Figure 3.**
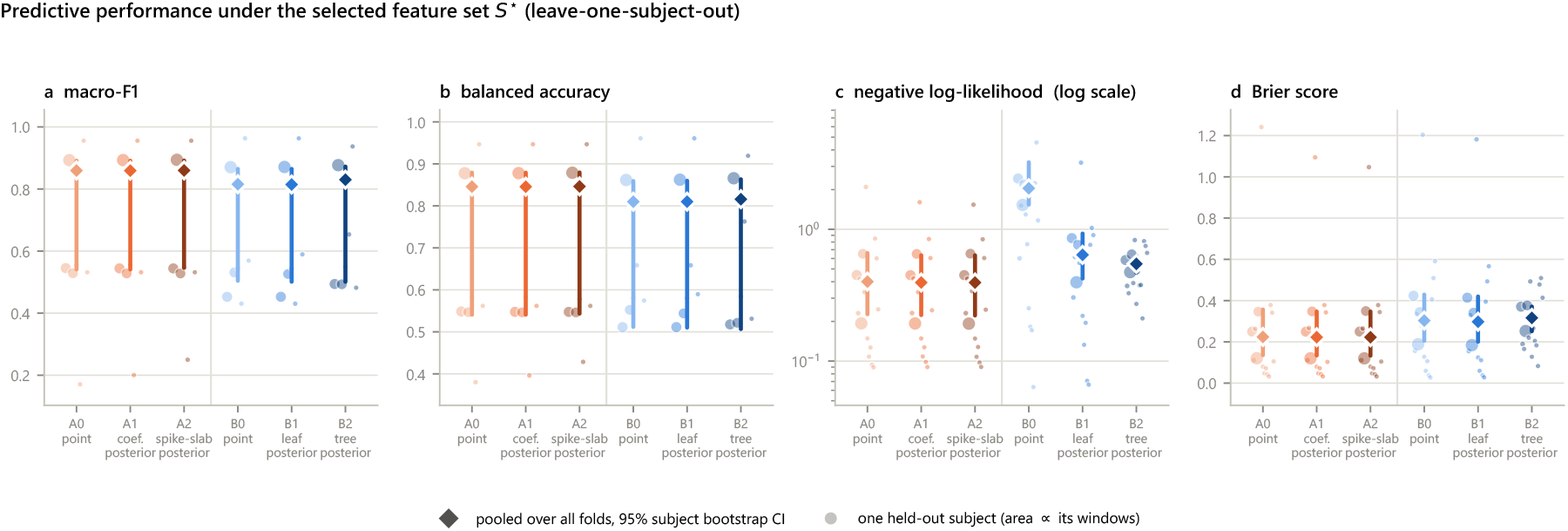
Predictive performance under *S*^⋆^ with leave-one-subject-out validation: (a) macro-F1, (b) balanced accuracy, (c) NLL and (d) Brier score. Diamonds are pooled estimates with 95% subject-bootstrap intervals; circles are held-out subjects.

The two families now sit at comparable operating points, which is what makes the uncertainty comparison interpretable. In the logistic family, macro-F1 is 0.859 (A0), 0.849 (A1) and 0.851 (A2), with NLL 0.401, 0.420 and 0.419 and ECE 0.040, 0.044 and 0.044. The paired A1−A0 contrast includes zero on every metric, so introducing the coefficient prior and its posterior does not measurably change what the linear model predicts. A2−A1 excludes zero on NLL (−0.0018 [−0.0051, −0.0002]) and Brier (−0.0012 [−0.0032, −0.0003]), but the effects are three orders of magnitude below the metrics they act on — intervals that exclude zero only at their own resolution.

In the tree family, macro-F1 is 0.815 (B0), 0.814 (B1) and 0.830 (B2). B1−B0 leaves discrimination untouched while repairing the likelihood: NLL falls from 2.045 to 0.640, a paired difference of−1.41 [−2.32, −1.11], with Brier −0.0057 [−0.0093, −0.0036], both excluding zero. An empirical leaf assigns probability zero to classes it did not observe, which is catastrophic under a log score; the Dirichlet posterior fixes that and nothing else. B2−B1 improves macro-F1 by 0.016 and NLL by 0.093, but every interval includes zero, so on 15 subjects the structure posterior cannot be separated from no change in predictive terms.

Q2 recall remains the weakest class recall for the linear rungs (0.642—0.681), the expected consequence of a boundary defined by whether morphology is interpretable rather than by noise amplitude.

#### Estimator checks

Numerical diagnostics establish that the posterior comparisons are not driven by failed optimisation or an obviously collapsed sampler. A0 converges to a unique maximum-likelihood solution with positive-definite curvature in every fold; A1 has positive-definite Laplace curvature and reproducible modes; A2 can be enumerated exactly over all 512 masks; and B2 retains 194—230 distinct tree structures per fold. These checks address whether the posterior calculations are numerically credible; they are distinct from the scientific question of *what* each posterior varies.

#### Posterlor objects and structural support

Figure 4 shows what varies at each posterior rung. Panel (a) shows the A1 posterior over the Q1—Q3 and Q2—Q3 coefficient contrasts. For *c* ∈ {Q1, Q2},

**Figure 4.**
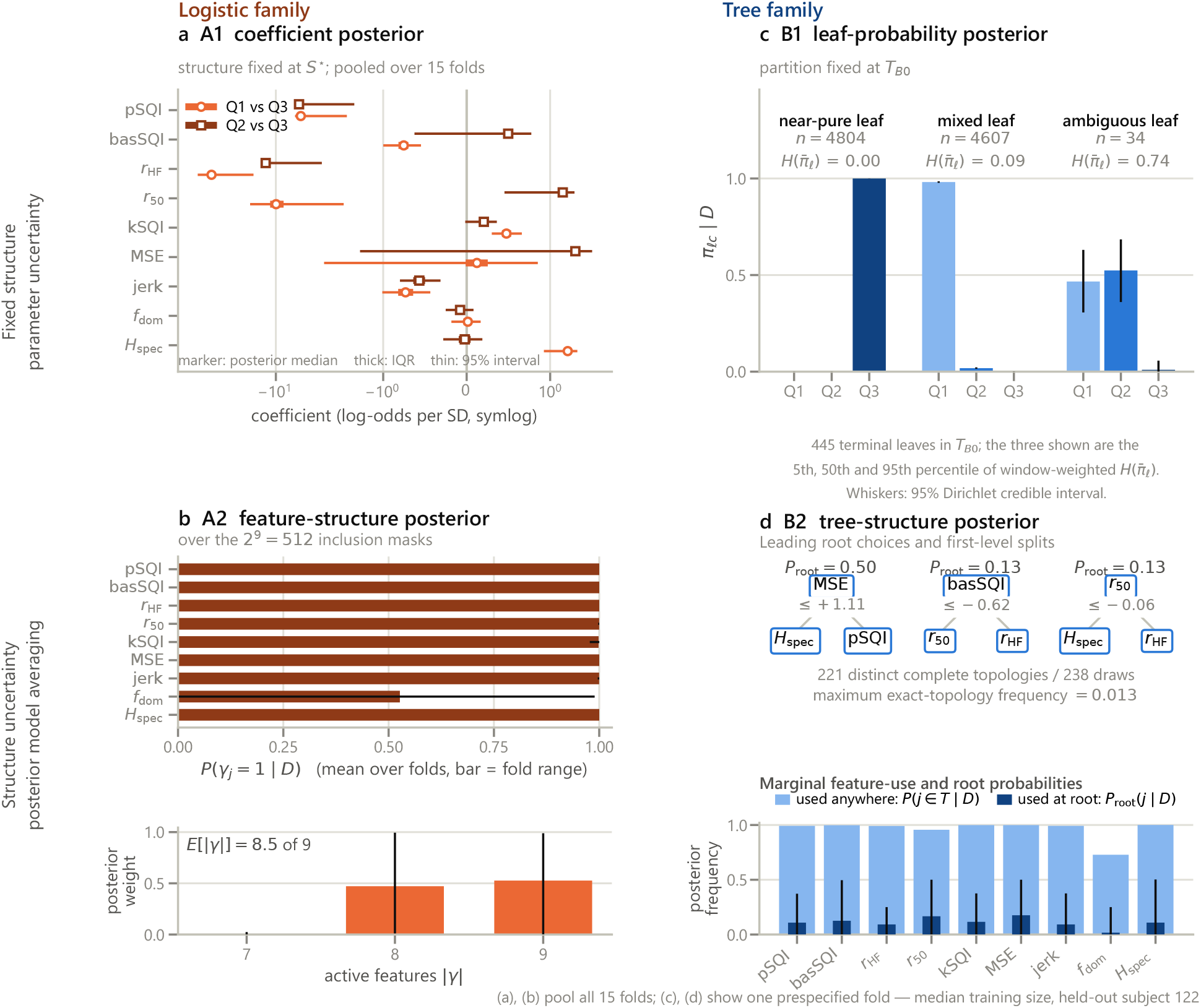
Posterior objects in the matched ladders: (a) A1 coefficient posterior; (b) A2 feature-inclusion and model-size posterior; (c) B1 leaf-probability posterior; and (d) B2 leading split structures with marginal feature-use and root probabilities. A1/A2 are fold summaries; B1/B2 use the prespecified reference fold.

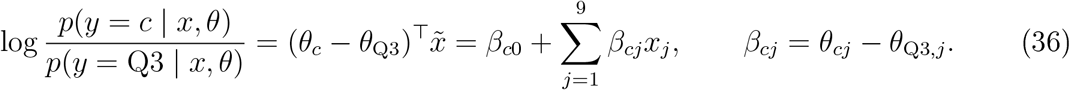

Because inputs are standardised, *β*_*cj*_ is the change in *c*-versus-Q3 log-odds for a one-standard-deviation increase in feature *j*; the symmetric-logarithmic axis in panel (a) is only a display scale. B1 is a posterior over class probabilities within the fixed B0 leaves. For A2, feature-structure support is summarised by the posterior inclusion probability (PIP),

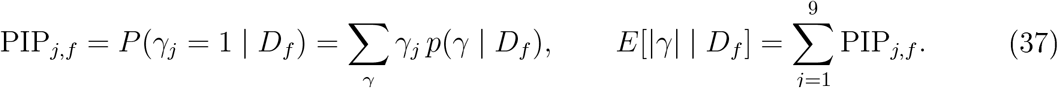

Across folds, eight features have mean PIP *>* 0.9, while *f*_dom_ has mean PIP ≃ 0.53 and *E*[|*γ*|] ≃ 8.5*/*9. A2 therefore averages mainly over near-complete feature masks, which explains the small change from A1.

For B1, representative leaves are summarised by

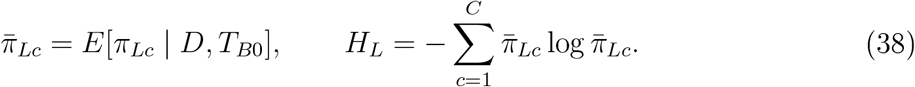

Panel (c) therefore contrasts near-pure, mixed and ambiguous leaves within one fixed partition. B2 instead places posterior mass over tree structures. Its marginal structural summaries are

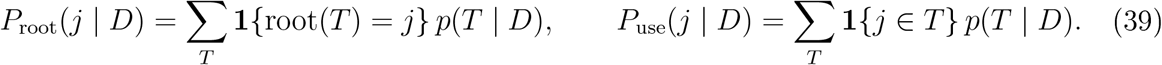

Figure 4d separates coarse from exact structure. The upper sketches show the leading root choices and their first-level splits, while the lower bars show the marginal probability that each feature is used anywhere in a tree or used specifically at the root. In the reference fold, MSE is the root in approximately 50% of draws, yet 221 distinct complete topologies occur among 238 retained draws and the most frequent topology has posterior frequency 0.013. Thus the selected predictors are repeatedly reused, but their ordering, thresholds and downstream partitioning remain uncertain. This structural diversity contributes to epistemic uncertainty only when the resulting trees give materially different predictions for the same observation.

### 3.3 Incremental reliability relative to max-softmax uncertainty

Figure 5 expresses the uncertainty results directly as paired gains over the confidence each model’s own predictive mean already supplies. This comparison is deliberately stricter than comparing rungs to one another: within each contrast the predicted probabilities, the predicted classes and therefore the errors are identical, and only the uncertainty ranking changes. Table 3 reports the same endpoints in absolute form.

**Table 3.** Reliability of posterior uncertainty against the same model’s *U*_MS_. Within each block the predictions are identical and only the uncertainty score changes, so differences reflect ranking quality rather than predictive accuracy. Lower AUGRC is better; higher error AUROC and *ρ* are better.

| score | error AUROC | AUGRC | $\rho(U, H_{\text{expert}})$ |
| --- | --- | --- | --- |
| *A1 – Laplace logistic* |  |  |  |
| $U_{\text{MS}}$ | 0.801<br>[0.665, 0.892] | 0.037<br>[0.014, 0.074] | 0.350<br>[0.026, 0.445] |
| $U_{\text{pred}}$ | 0.797<br>[0.662, 0.889] | 0.037<br>[0.014, 0.075] | 0.353<br>[0.022, 0.444] |
| $U_{\text{CE}}$ | 0.793<br>[0.629, 0.887] | 0.038<br>[0.014, 0.080] | 0.353<br>[0.035, 0.443] |
| $U_{\text{MI}}$ | 0.776<br>[0.679, 0.862] | 0.040<br>[0.017, 0.086] | 0.406<br>[0.072, 0.457] |
| *A2 – spike-and-slab logistic* |  |  |  |
| $U_{\text{MS}}$ | 0.801<br>[0.667, 0.891] | 0.037<br>[0.014, 0.074] | 0.347<br>[0.025, 0.445] |
| $U_{\text{pred}}$ | 0.797<br>[0.663, 0.889] | 0.037<br>[0.014, 0.074] | 0.350<br>[0.021, 0.444] |
| $U_{\text{CE}}$ | 0.793<br>[0.631, 0.887] | 0.038<br>[0.014, 0.078] | 0.351<br>[0.036, 0.443] |
| $U_{\text{MI}}$ | 0.757<br>[0.672, 0.826] | 0.042<br>[0.020, 0.087] | 0.370<br>[0.069, 0.446] |
| *B1 – Dirichlet-leaf CART* |  |  |  |
| $U_{\text{MS}}$ | 0.774<br>[0.675, 0.813] | 0.060<br>[0.027, 0.113] | 0.269<br>[0.032, 0.374] |
| $U_{\text{pred}}$ | 0.767<br>[0.678, 0.811] | 0.061<br>[0.027, 0.119] | 0.277<br>[0.058, 0.375] |
| $U_{\text{CE}}$ | 0.767<br>[0.673, 0.805] | 0.061<br>[0.028, 0.116] | 0.266<br>[0.043, 0.374] |
| $U_{\text{MI}}$ | 0.749<br>[0.653, 0.851] | 0.064<br>[0.023, 0.127] | 0.326<br>[0.128, 0.369] |
| *B2 – Bayesian CART* |  |  |  |
| $U_{\text{MS}}$ | 0.633<br>[0.614, 0.774] | 0.072<br>[0.038, 0.098] | 0.054<br>[-0.018, 0.109] |
| $U_{\text{pred}}$ | 0.572<br>[0.519, 0.768] | 0.081<br>[0.048, 0.103] | -0.003<br>[-0.051, 0.099] |
| $U_{\text{CE}}$ | 0.685<br>[0.571, 0.830] | 0.064<br>[0.022, 0.111] | 0.254<br>[0.047, 0.386] |
| $U_{\text{MI}}$ | 0.469<br>[0.366, 0.705] | 0.097<br>[0.059, 0.129] | -0.137<br>[-0.220, 0.025] |

**Figure 5.**
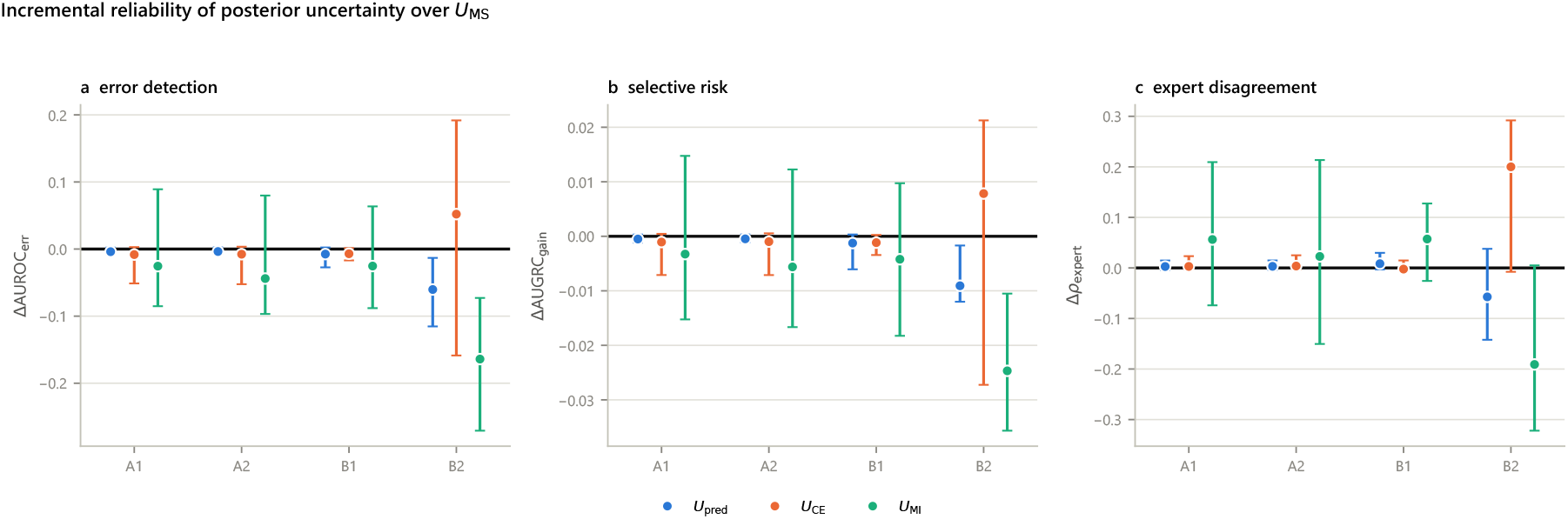
Incremental reliability relative to same-model max-softmax uncertainty *U*_MS_: (a) error-detection AUROC gain, (b) AUGRC gain and (c) gain in association with expert vote entropy. Positive values favour the posterior-derived score; intervals are 95% subject-bootstrap confidence intervals.

#### Logistic-family comparison

For both A1 and A2, *U*_pred_ and *U*_CE_ sit fractionally *below* the zero line on every endpoint. At A2, ΔAUROC_err_(*U*_pred_) = −0.004 [−0.007, 0.002] and ΔAUGRC_gain_(*U*_pred_) = −0.0005 [−0.0011, 0.0003]; the corresponding figures at A1 are −0.004 and −0.0005. In absolute terms the three scores are the same number to two decimal places — error AUROC 0.797 for *U*_pred_ against 0.801 for *U*_MS_, AUGRC 0.037 for both. Introducing coefficient uncertainty at A1 and beta-binomial spike-and-slab structure uncertainty at A2 therefore changes the posterior representation without producing a corresponding gain in reliability over the probability baseline, even though the A2 structure posterior is not concentrated on one structure (Section 3.2).

#### Mutual-information behaviour

*U*_MI_ is the weakest error ranker anywhere in the logistic family — ΔAUROC_err_ = −0.026 at A1 and −0.044 at A2, absolute AUROC 0.776 and 0.757 against 0.801 for *U*_MS_ — and yet it has the strongest association with annotator disagreement in that family, *ρ* = 0.406 at A1 against 0.350 for *U*_MS_ (Δ*ρ*_expert_ = +0.056, [−0.074, 0.209]). The same dissociation appears at B1, where *U*_MI_ again ranks errors worst and correlates with annotator disagreement best (Δ*ρ*_expert_ = +0.057).

That is worth stating carefully, because it cuts against the simple reading in both directions. The epistemic component is not uninformative; it is informative about a different aspect of reliability. Windows on which the posterior disagrees with itself are disproportionately windows on which the three human annotators also disagreed, but they are not the windows on which the model is most likely to be wrong. A gate built on *U*_MI_ would defer ambiguous recordings rather than incorrect ones, and those are not the same operating policy. The intervals on Δ*ρ*_expert_ are wide enough that this is a direction, not a demonstration.

#### Tree-family comparison

At B1 all three posterior scores are marginally below *U*_MS_ and none is distinguishable from it. Adding the tree-structure posterior at B2 reorders them: *U*_CE_ becomes the only score in the study with a positive point estimate on every endpoint (ΔAUROC_err_ = +0.052, ΔAUGRC_gain_ = +0.008, Δ*ρ*_expert_ = +0.200), while *U*_pred_ falls to −0.060 [−0.115, −0.013] and *U*_MI_ to −0.164 [−0.271, −0.073] on error AUROC. Between two rungs of the same family, fitted on the same nine columns, the most informative decomposition term changes from *U*_MI_ to *U*_CE_ and the least informative changes from *U*_CE_ to *U*_MI_. The direction of any incremental gain therefore depends on where posterior variability is represented, not on the presence of a Bayesian posterior as such — and even B2’s *U*_CE_ advantage has an interval spanning zero on all three endpoints, so it is a candidate rather than a result.

#### Association with annotator disagreement

The largest absolute correlation in Table 3 is 0.406 and several intervals include zero, even though the ambiguity is real: the three annotators split on 8,320 of the 32,224 windows (25.8%). Posterior uncertainty is a model-dependent diagnostic whose value has to be demonstrated for the intended decision criterion, not an intrinsically superior confidence score, and it is not a substitute for a measurement of human ambiguity.

#### Between-rung comparison

The comparisons above hold the model fixed and vary the score. When the model itself moves up a rung, both the predictions and the uncertainty score can change, so differences in selective risk can reflect predictive accuracy as well as ranking quality. Within the logistic family the operating curves change little, consistent with the flat contrasts of Section 3.1. Within the tree family, the B0-to-B2 change is more substantial because the predictions and confidence profile both change; it should therefore not be read as a pure uncertainty-ranking effect.

### 3.4 Behaviour of aleatoric and epistemic uncertainty

Section 2.5 designates *U*_CE_ as the aleatoric/data component and *U*_MI_ as the epistemic/model component. Figure 6 tests these roles; the expected directions are present, but the responses are not fully component-specific.

**Figure 6.**
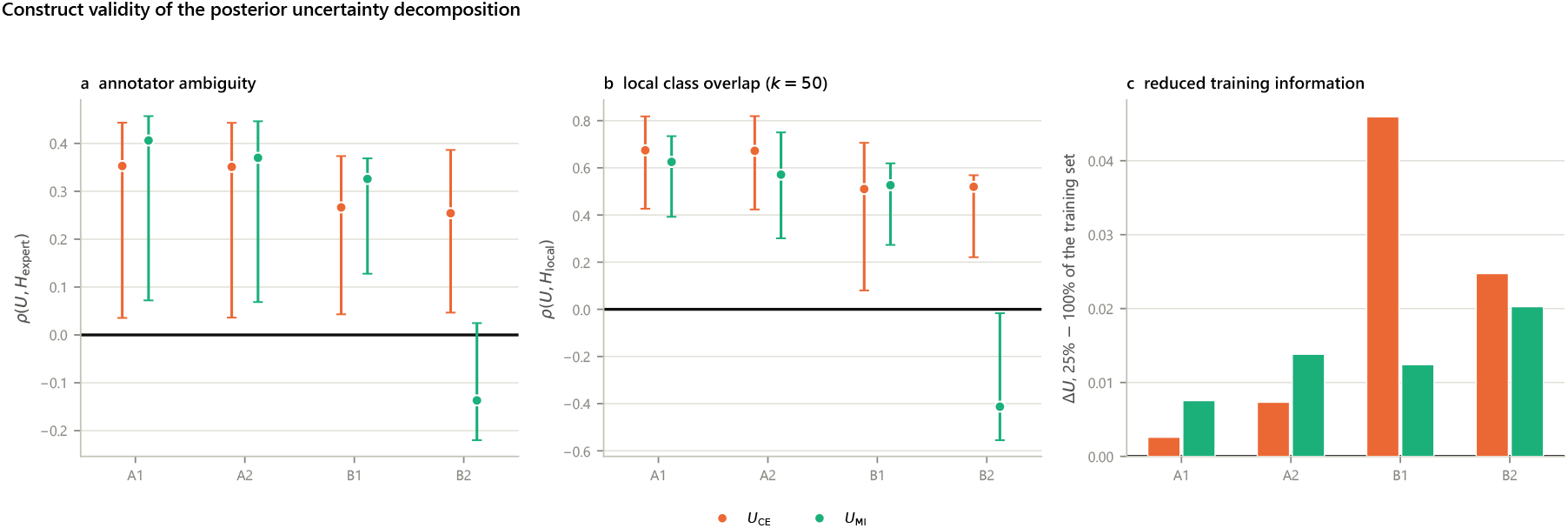
Behavioural probes for the designated uncertainty components: (a) expert-vote entropy, (b) local class-overlap entropy and (c) reduced training information. Intervals in (a) and (b) are 95% subject-bootstrap confidence intervals.

#### Local class overlap

For each held-out window, *H*_local_ is the label entropy of its 50 nearest training neighbours in the locked standardised feature space. Sixty-nine per cent of held-out windows lie in a mixed-label neighbourhood. The correlation with *H*_local_ is stronger for *U*_CE_ than for *U*_MI_ at three of four posterior rungs: 0.674 versus 0.625 at A1, 0.672 versus 0.571 at A2, and 0.520 versus −0.413 at B2; B1 is a near-tie in the opposite direction (0.510 versus 0.526). At A2, *ρ*(*U*_CE_, *H*_local_) = 0.672 [0.423, 0.819], the strongest construct association in the study. This supports the assigned aleatoric role of *U*_CE_ when ambiguity is defined as local class overlap in the available representation.

#### Expert disagreement

Correlations with expert vote entropy are weaker overall and reverse the previous ordering at A1, A2 and B1: 0.406 versus 0.353 at A1, 0.370 versus 0.351 at A2, and 0.326 versus 0.266 at B1 for *U*_MI_ versus *U*_CE_. At B2, *U*_CE_ is larger while *U*_MI_ falls to −0.137. Thus local class overlap and human disagreement are not interchangeable ambiguity proxies. *U*_CE_ captures the former more consistently, whereas posterior disagreement can align more closely with the latter.

#### Reduced training information

Refitting on one quarter of each training fold increases *U*_MI_ at every posterior rung: by 0.008 at A1, 0.014 at A2, 0.012 at B1 and 0.020 at B2. This is the expected direction for the designated epistemic component. However, *U*_CE_ also increases, and at both tree rungs its change is larger: 0.046 versus 0.012 at B1 and 0.025 versus 0.020 at B2. The model-information manipulation therefore supports the epistemic interpretation directionally while also demonstrating cross-sensitivity.

Taken together, the probes show partial construct validity. *U*_CE_ behaves as an aleatoric component under local class overlap, and *U*_MI_ behaves as an epistemic component when training information is reduced, but neither component is isolated from the other’s manipulation. Their operational meaning therefore remains conditional on the model, representation and ambiguity proxy used.

### 3.5 Selective risk across uncertainty scores

Figure 7 shows the operational consequence of the same-model comparisons for the richest posterior rung of each family. Windows are accepted from lowest to highest uncertainty, so a useful score keeps selective risk low as coverage increases. Because all curves within a panel rank the predictions of one fixed model, they necessarily meet at full coverage; differences before that point are therefore differences in ranking rather than differences in predictive accuracy. The main figure focuses on the two structure-posterior rungs, which provide the clearest comparison of uncertainty ranking after structural uncertainty is introduced.

**Figure 7.**
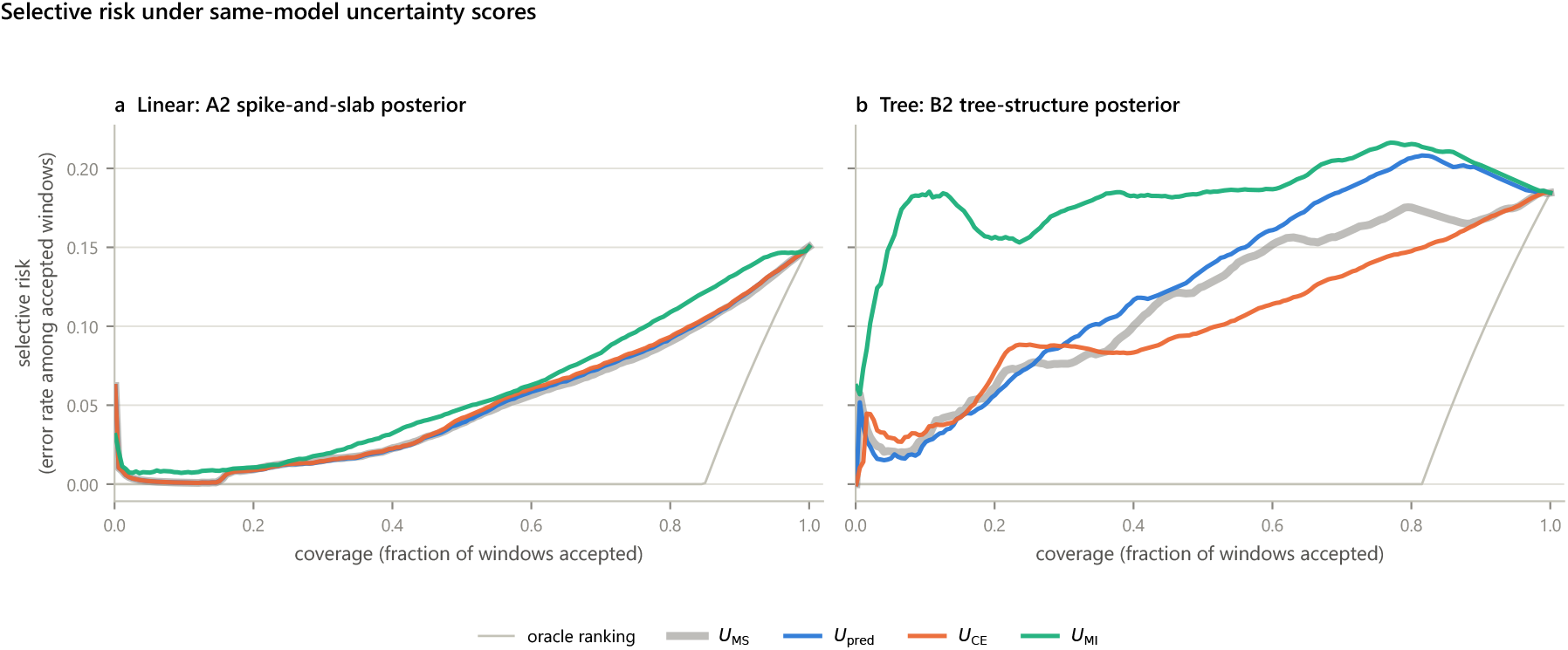
Selective risk for the structure-posterior rungs A2 and B2. Curves compare *U*_MS_, *U*_pred_, *U*_CE_ and *U*_MI_ on identical predictions; lower risk at a given coverage is better. Grey denotes the oracle ranking.

A2 exhibits a markedly more stable uncertainty—risk ordering than B2. Across most of the coverage range *U*_MS_, *U*_pred_ and *U*_CE_ trace nearly overlapping trajectories, and progressively restricting acceptance to lower-uncertainty windows produces a smooth reduction in retained-set error: rejecting the least certain half takes selective risk from 0.162 at full coverage to 0.048, for the posterior scores and for *U*_MS_ alike. Only *U*_MI_ is separated from the others, and consistently so. A deployment could pick any of the first three and obtain the same policy.

B2 is substantially more score dependent. *U*_CE_ holds the lowest selective risk over much of the operating range, while *U*_MI_ retains a high error rate even at low coverage, sitting far above the others from roughly 5% coverage upward. On the same fixed predictions, two components of one decomposition therefore imply different rejection sets.

Posterior complexity alone does not determine selective utility. A2 carries the richer linear posterior and gains nothing from it over *U*_MS_; what it does offer is a mapping from uncertainty score to realised risk that is insensitive to which score is chosen, which is precisely what a threshold-based policy needs. B2 offers a component that beats its own baseline and a component that is much worse than it, with no way to tell them apart from in-distribution calibration.

### 3.6 Uncertainty under controlled ECG corruption

Figure 8 adds independently recorded NSTDB artifact to held-out BUT QDB ECG at controlled SNR. Because the artifacts come from different volunteers and Holter hardware/electrode placements, the perturbation includes acquisition heterogeneity while the underlying ECG remains fixed. Models remain frozen, and accuracy is not scored against clean labels because added artifact may genuinely change the signal-quality class. The analysis therefore separates predicted degradation from the uncertainty response.

**Figure 8.**
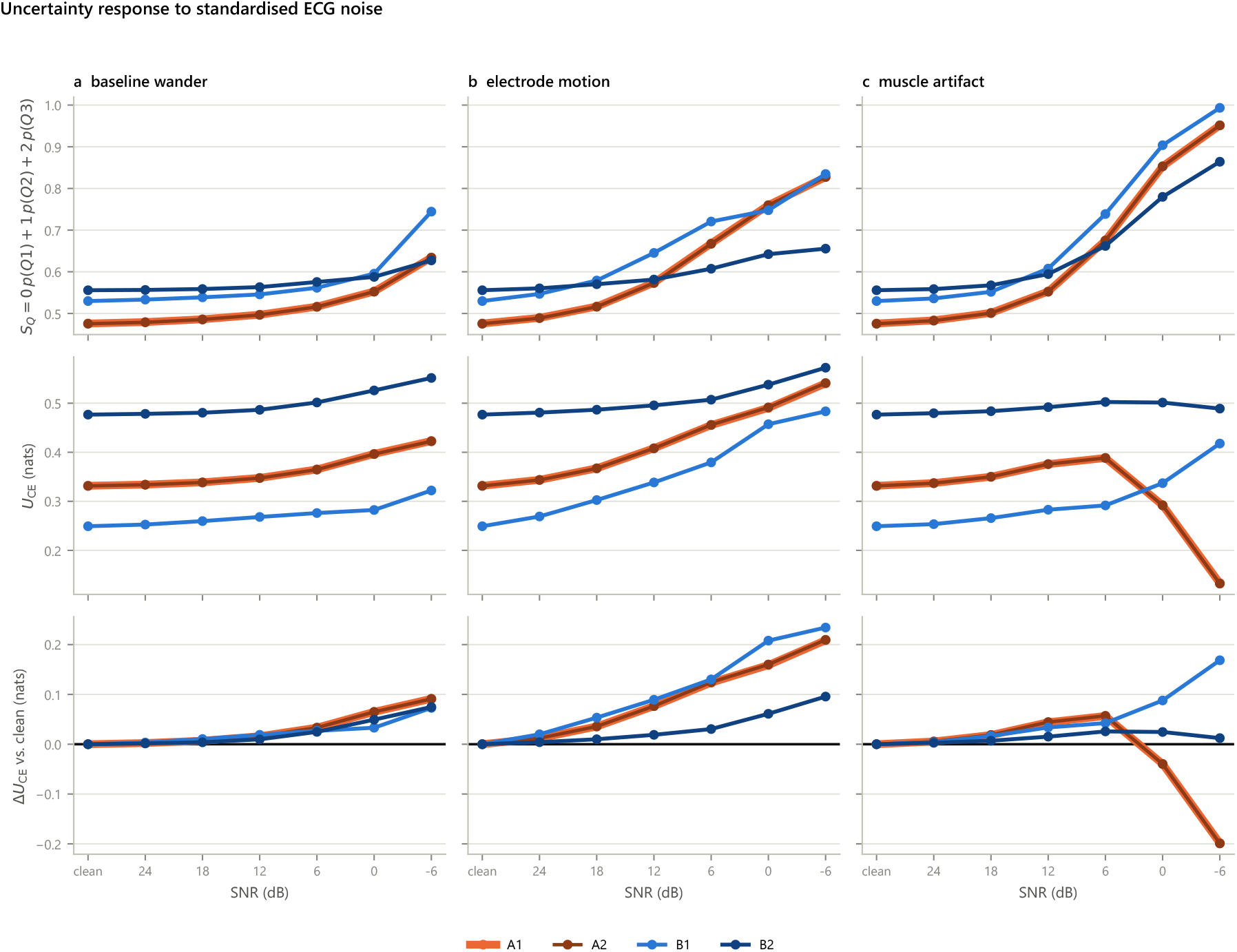
NSTDB stress test under baseline wander, electrode motion and muscle artifact. Rows show predicted severity *S*_Q_, aleatoric uncertainty *U*_CE_ and its change from the clean baseline. Only ECG is corrupted; models remain frozen.

#### Predicted quality severity

Predicted severity rises monotonically with contamination at every rung. At A2, *S*_*Q*_ increases from 0.476 on clean windows to 0.634, 0.827 and 0.951 at −6 dB under baseline wander, electrode motion and muscle artifact, respectively (Figure 8, top row). All six rungs reproduce this artifact ordering. Baseline wander mainly displaces the low-frequency baseline while preserving QRS structure, whereas muscle artifact more strongly obscures morphology; the frozen estimators therefore register the expected degradation pat-tern.

#### Uncertainty response

The middle row shows *U*_CE_ and the bottom row its change from each model’s clean baseline, Δ*U*_CE_(SNR) = *U*_CE_(SNR) − *U*_CE_(clean). Under baseline wander and electrode motion, *U*_CE_ rises with severity at every rung. At −6 dB, the increases over clean are +0.091 and +0.210 nats at A2, +0.073 and +0.234 at B1, and +0.075 and +0.096 at B2, with electrode motion producing the larger increase throughout. For these two artifacts, degradation and predictive ambiguity therefore increase together.

Muscle artifact separates the families. The linear rungs rise to +0.057 nats at +6 dB and then invert, falling to −0.039 at 0 dB and −0.199 at −6 dB, where *U*_CE_ = 0.13 nats, the lowest value observed in the stress test. The tree rungs do not invert: B1 reaches +0.169 and B2 remains near baseline at +0.012. A1 and A2 coincide throughout, so marginalising over active-feature structure does not alter the pattern.

#### Severity versus ambiguity

Muscle artifact separates predicted severity from predictive ambiguity. At +6 dB, the linear rungs show *S*_*Q*_ = 0.675 with elevated *U*_CE_, consistent with a borderline Q2—Q3 state. By −6 dB, *S*_*Q*_ reaches 0.951 while *U*_CE_ falls below clean, indicating a confidently poor-quality prediction rather than increasing ambiguity. The drop should therefore not be read as failure of uncertainty itself: once morphology is sufficiently degraded, the model can become more certain of a poor-quality state even while signal quality worsens. Severity and ambiguity need not be monotone in one another. This behaviour is compatible with a data-uncertainty term on a task whose target is signal quality, but correctness cannot be established without re-annotating the corrupted windows.

Operationally, the non-monotone response means *U*_CE_ cannot serve as a generic shift detector on this axis: a thresholded logistic gate could flag intermediate contamination yet miss the most severe inputs. The two families therefore encode the same perturbation differently rather than one being intrinsically correct: B1 and B2 continue to rise, consistent with the bounded regions of a tree partition and B2’s generally high entropy. Repeating the test with ECG-only 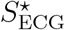 reproduces and slightly deepens the inversion, excluding the untouched accelerometer as its cause.

### 3.7 Zero-shot external transfer to LTSTDB

Adding recorded artifact at a controlled SNR fixes the direction and severity of the shift, but cannot reproduce the full differences of an independent database. Zero-shot transfer therefore evaluates the frozen BUT QDB models on LTSTDB (Jager et al., 2003; Goldberger et al., 2000), with no target-database fitting. This transfer changes several factors simultaneously: the subject cohort, recording sites and acquisition systems, lead configuration and digitisation, availability of accelerometry, and annotation protocol. BUT QDB’s three-class taxonomy and LTSTDB’s unreadable annotations were also developed independently (Nemcova et al., 2020). The external states are therefore the occupancy states defined in Eq. (34), with transition constructed from partial window overlap rather than supplied as an LTSTDB class. The question is whether the frozen BUT QDB severity score remains ordinal across these states, not whether Q1—Q3 accuracy is preserved across databases.

The transferred severity score is ordinal across the three occupancy states for all six rungs. *S*_*Q*_ rises from clean through transition to fully unreadable in every case: from 0.35 to 0.50 to 0.81 in the logistic family, from 0.70 to 1.00 to 1.40 at B0 and B1, and from 0.47 to 0.51 to 0.67 at B2 (Figure 9a). The ordering also holds within subject, where the transition-minus-clean and unreadable-minus-clean steps are positive at every rung (median +0.22 and +0.24 in the logistic family), while the unreadable-minus-transition step is small and, at B2, marginally negative (Figure 9b). Descriptively, a model trained on one site’s three-level wearable taxonomy assigns higher ordinal severity to a different site’s independently annotated signal failure without any target-site fitting. Given that only seven LTSTDB subjects contain duration-bounded unreadable intervals, this is transfer evidence rather than population-level validation.

**Figure 9.**
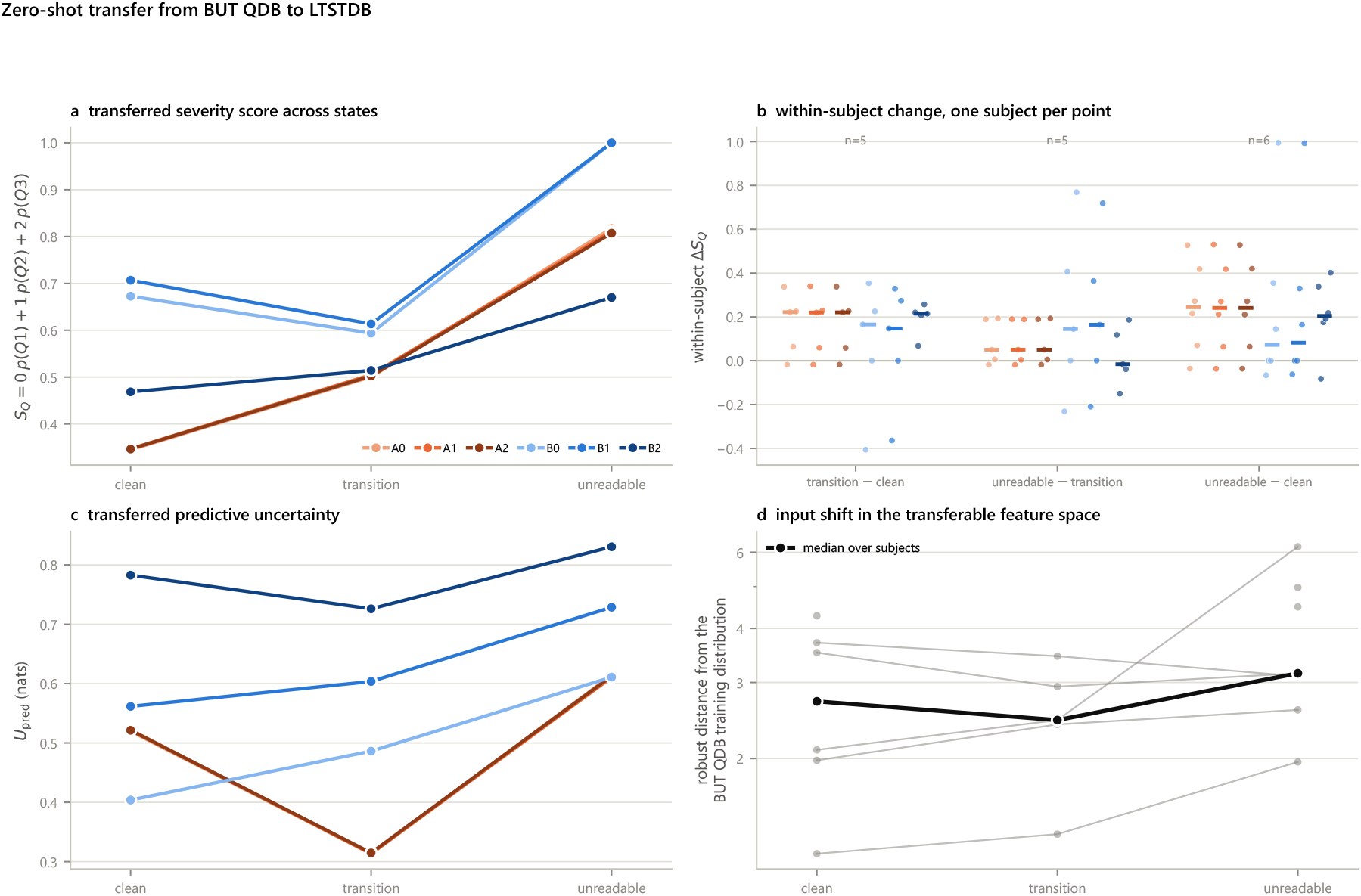
Zero-shot BUT QDB→LTSTDB transfer: (a) ordinal severity S_Q_, (b) within-subject severity steps, (c) predictive entropy and (d) robust feature-space distance from BUT QDB. LTSTDB occupancy states are defined in Eq. (34).

Transferred uncertainty does not follow the same pattern, and that is the substantive result of the section. No rung reproduces the monotone progression its own severity score shows. The linear rungs are non-monotone in the direction least useful for a gate: *U*_pred_ falls from 0.52 on clean windows to 0.31 on transition windows before recovering to 0.61 on fully unreadable ones, so the state in which the window is only *partly* corrupted — the ambiguous case — is the one on which the frozen linear models are most confident. B1 peaks on transition windows (0.61 against 0.55 clean and 0.51 unreadable), and B0 is highest on clean windows. Only B2 is both high throughout and highest on unreadable windows (0.83 against 0.78 clean), and even it dips on the transition state (Figure 9c). Prediction transfers more consistently than uncertainty does, and the two would give a deployed gate different instructions.

Figure 9d asks whether the inputs themselves moved, in the same six transferable columns and against the same three states. Measured as a robust distance from the BUT QDB training median, LTSTDB windows sit two to three robust standard deviations away even when LT-STDB calls them clean, and the fully unreadable windows sit farthest of all: the median over subjects is 2.75 on clean, 2.50 on transition and 3.18 on unreadable. The progression is not uniform — the transition state is slightly closer than clean at the median, and individual subjects move in both directions — but the endpoints are ordered and the shift is unambiguously present.

That is the connection back to Section 3.6. The input moves away from the training distribution as the annotated state worsens, and the transferred severity score responds, yet predictive uncertainty does not track the shift proportionally. Unlike the controlled NSTDB mixture, however, LTSTDB bundles population and acquisition changes with the label-definition shift. The result therefore demonstrates robustness to a composite external domain change rather than isolating which population, instrumentation, recording protocol, or annotation factor caused the uncertainty behaviour.

## 4 Discussion

The experiments were designed to test uncertainty as an observable reliability construct rather than to compare Bayesian and non-Bayesian models solely by predictive accuracy. Three methodological patterns recur across the results. First, posterior enrichment is useful only when it creates decision-relevant predictive diversity. Second, conventional aleatoric/epistemic labels require behavioural validation because their corresponding scores are not source-pure. Third, distribution shift can alter predicted signal-quality severity without producing a monotonic increase in model uncertainty. These findings argue against treating posterior complexity, entropy magnitude or Bayesian formulation alone as evidence that an uncertainty estimate is operationally trustworthy.

### 4.1 Posterior richness is not equivalent to reliability

The matched model ladders show that a richer posterior is useful only when its plausible models disagree in prediction. A1 introduces coefficient uncertainty while keeping all nine features fixed. A2 adds feature-structure uncertainty, but Figure 4b shows that the posterior is concentrated on near-complete masks: eight features have mean inclusion probability above 0.9 and *E*[|*γ*|] ≃ 8.5*/*9. The selected representation is therefore structurally stable within this cohort and logistic family, with most uncertainty concentrated on *f*_dom_. Because the supported masks retain nearly the same information, A2 adds little predictive disagreement beyond A1 and correspondingly little change in uncertainty reliability.

B2 exposes a different form of structure uncertainty. B1 keeps the B0 partition fixed and updates only the leaf probabilities, whereas B2 averages over alternative partitions. In the reference fold, 221 distinct complete topologies occur among 238 retained draws, although coarse preferences remain: MSE is the root split in about 50% of draws. The posterior is therefore not uncertain about whether the selected predictors are useful; it is uncertain about how they should be organised into split variables, thresholds and downstream branches. This creates substantially more opportunity for predictive disagreement than omitting one weak feature from an otherwise stable logistic model, which explains the stronger change in uncertainty ordering from B1 to B2.

Structural diversity should nevertheless not be equated directly with epistemic uncertainty. Different posterior-supported models may still assign similar class probabilities to the same window; *U*_MI_ increases only where their predictions diverge. Posterior diagnostics and operational usefulness are therefore separate questions: curvature, prior-sensitivity and MCMC checks establish computational credibility, whereas reliability depends on whether posterior diversity is informative about failure. The same distinction applies to ensemble uncertainty quantification (Lakshminarayanan et al., 2017; Ovadia et al., 2019).

### 4.2 Interpreting aleatoric and epistemic uncertainty

The behavioural probes support the use of *U*_CE_ as the aleatoric/data component and *U*_MI_ as the epistemic/model component, while also showing why post-hoc entropy decomposition should not be read as complete physical source separation. The strongest convergent result is the association between *U*_CE_ and local class overlap: at A2, *ρ*(*U*_CE_, *H*_local_) = 0.672. This matches the conditional-entropy interpretation because these windows occupy regions of the selected representation where training labels overlap.

The complementary epistemic manipulation also behaves in the expected direction: reducing each training fold to one quarter increases *U*_MI_ at every posterior rung. The response is not specific, however, because *U*_CE_ also changes, particularly in the tree family. Expert-vote entropy provides a second illustration. *U*_MI_ is more strongly associated with expert disagreement at A1, A2 and B1, even though it is generally the weaker error-ranking score. Human disagreement, local overlap and model disagreement therefore capture related but different forms of ambiguity.

This pattern is consistent with prior analyses showing that conditional entropy and mutual information depend on the fitted model, posterior approximation and available representation (Hüllermeier and Waegeman, 2021; Wimmer et al., 2023; Gruber et al., 2023). We retain the conventional aleatoric/epistemic terminology because the prespecified manipulations support those roles directionally. The important qualification is construct specificity: *U*_CE_ and *U*_MI_ are model-based uncertainty components whose meanings are validated by behaviour, not ground-truth measurements of two independent physical sources.

### 4.3 Selective decisions depend on score and threshold

The same-model risk—coverage analysis turns those statistical differences into an operational question: which windows would a thresholded system actually retain or reject? Because the predictions are fixed within each panel, differences between *U*_MS_, *U*_pred_, *U*_CE_ and *U*_MI_ are differences in the rejection order alone (El-Yaniv and Wiener, 2010; Geifman and El-Yaniv, 2017; Geifman et al., 2019; Traub et al., 2024). A2 is stable in this sense: *U*_MS_, *U*_pred_ and *U*_CE_ generate almost the same risk—coverage trajectory. That stability is useful for threshold selection, but it should not be mistaken for a Bayesian gain because the posterior-derived scores do not improve on *U*_MS_.

B2 presents the opposite trade-off. Its *U*_CE_ gives the best selective ordering over much of the coverage range, whereas *U*_MI_ can retain a high error rate even after many predictions are rejected. Thus two components of the same exact entropy decomposition can imply materially different actions on identical predictions. The important deployment object is consequently the triplet of model, uncertainty score and operating threshold, not the uncertainty label in isolation.

The disagreement result sharpens this point. *U*_MI_ can identify windows that the experts also find ambiguous without identifying the windows on which the model is most likely to be wrong. A system optimised for human referral, for example, may value that behaviour differently from a system whose objective is to minimise silent classification failures. Reliability therefore has to be defined against the downstream decision being supported before an uncertainty score is selected.

### 4.4 Uncertainty under controlled and external distribution shift

The shift experiments show why in-distribution selective performance is insufficient for wearable deployment. Under NSTDB contamination, predicted quality severity rises monotonically for baseline wander, electrode motion and muscle artifact, whereas *U*_CE_ depends on the ambiguity regime. In particular, severe muscle artifact can move the logistic models from an ambiguous intermediate state to a confident poor-quality decision. Signal degradation and ambiguity about the resulting class are therefore distinct quantities, and posterior uncertainty should not be treated as a generic distribution-shift detector.

The acquisition context also matters. NSTDB contributes artifact recorded from different volunteers with Holter hardware and electrode placement, but the underlying test ECG remains a BUT QDB Bittium-Faros recording; it is therefore a controlled perturbation with only partial acquisition heterogeneity. LTSTDB is a stronger composite shift: it changes cohort, clinical sites, ambulatory recording systems, lead configuration and digitisation, removes the accelerometer modality, and uses an independently defined annotation scheme. Lead configuration is not even held fixed inside the target: the annotated channel is precordial in some records, bipolar limb-type in others and orthogonal E-S in the rest, so the shift is heterogeneous across LTSTDB subjects as well as between databases. Its severity ordering transfers more consistently than its uncertainty ordering even though feature-space distance confirms substantial domain movement.

These results do not identify which source of shift dominates the uncertainty response. A deployment study should therefore distinguish population shift from instrumentation, lead placement, recording protocol and annotation shift where possible. Model-derived uncertainty may also need to be paired with an input-domain or anomaly score that responds directly to unfamiliar observations (Ibrahim et al., 2026; Malinin and Gales, 2018; Moon et al., 2020).

### 4.5 Limitations and future validation

Several modelling choices deliberately bound the present conclusions. First, the signal features are a compact set of interest chosen for interpretability and prior use in ECG-quality and wearable-motion studies, not an exhaustive representation of all possible ECG—accelerometer information (Clifford et al., 2012; Orphanidou et al., 2015; Zhao and Zhang, 2018; Beach et al., 2021; Hamidi et al., 2023). The screening result supports E3+E4+M2 in this cohort, and A2 further shows that eight of its nine features are retained with high posterior probability. M3 remains literature-grounded; its exclusion means only that the adapted coupling summaries were conditionally redundant once spectral, complexity and motion-dynamics features were present. Learned multimodal representations may expose different uncertainty behaviour and should be tested on larger cohorts.

Second, the uncertainty expressions are tied to categorical softmax prediction. Max-softmax uncertainty, predictive entropy and the conditional-entropy/mutual-information decomposition are convenient because they operate on the same class-probability vector and are widely used in classification UQ. Softmax probability, however, is not uniquely optimal for expressing uncertainty and can remain overconfident under covariate or out-of-distribution shift; alternative output parameterisations can behave differently (Padhy et al., 2020; Ovadia et al., 2019). A regression task would require a different predictive object, such as a posterior predictive density, variance, quantiles or intervals, together with regression-specific reliability metrics rather than max-softmax or class entropy (Bench et al., 2026; de Jong et al., 2026).

The empirical scope is also limited by the 15-subject BUT QDB cohort and its subject imbalance. The representation screen is not nested inside the outer LOSO folds, so absolute performance conditional on *S*^⋆^ is post-selection; LTSTDB provides a composite external shift rather than a replication of the three-class problem. The Laplace approximation in A1/A2 and finite B2 MCMC sampling introduce additional model-approximation error, and the NSTDB experiment alters ECG without physically coupled accelerometer artifact. Future studies should therefore use larger multi-site ECG—accelerometer cohorts and cross-platform acquisition to separate uncertainty associated with cohort composition, recording hardware, lead placement, sampling and acquisition protocols from uncertainty associated with the predictive model itself. Nested representation assessment, richer posterior or ensemble families, and prospective uncertainty-aware abstention, reacquisition or decision policies should then be evaluated under these controlled shifts. Such policies include JITAI-like systems in which an upstream signal-state estimate is gated by a validated reliability signal before adaptive action (Nahum-Shani et al., 2018; Liao et al., 2020).

## 5 Conclusion

This study used wearable ECG signal-quality assessment as a methodological test bed for asking when predictive uncertainty can be treated as a reliability signal. Across matched logistic-regression and CART families, posterior enrichment did not have a uniform effect. In the logistic family, A2 retained an expected 8.5 of nine selected features and concentrated support on near-complete masks, so the additional structural posterior produced little new predictive diversity. B2 instead averaged over materially different CART partitions, producing greater structural and predictive disagreement and a stronger dependence of selective-risk behaviour on the uncertainty score.

The behavioural experiments also show why uncertainty labels should be validated rather than assumed. Expected conditional entropy responded strongly to local class overlap, while mutual information increased when training information was reduced, supporting their designated aleatoric and epistemic roles directionally. Their cross-sensitivity, however, indicates that they are model-dependent constructs rather than direct measurements of independent physical sources of uncertainty. Under recorded ECG corruption and zero-shot LTSTDB transfer, predicted signal-quality severity changed more consistently than uncertainty ordering, showing that degradation, unfamiliarity and model uncertainty are related but non-equivalent quantities.

The main methodological conclusion is therefore that Bayesian formulation and posterior complexity do not by themselves establish trustworthy uncertainty. Reliability must be demonstrated against the intended use of the score, including error ranking, selective rejection, ambiguity sensitivity, information sensitivity and behaviour under relevant distribution shifts. For wearable ECG systems, future multi-site studies should test these criteria across acquisition hardware, lead configurations, cohorts and annotation protocols before uncertainty is used to gate abstention, reacquisition or human review.

## Data Availability

All data used in this study are publicly available online through PhysioNet: the Brno University of Technology ECG Quality Database (BUT QDB) at https://physionet.org/content/butqdb/1.0.0/, the MIT-BIH Noise Stress Test Database (NSTDB) at https://physionet.org/content/nstdb/1.0.0/, and the Long-Term ST Database (LTSTDB) at https://physionet.org/content/ltstdb/1.0.0/.

https://physionet.org/content/nstdb/1.0.0/

https://physionet.org/content/ltstdb/1.0.0/

## Data availability statement

BUT QDB, the MIT-BIH Noise Stress Test Database (NSTDB), and LTSTDB are publicly available from PhysioNet. The analysis code used to produce the figures and tables in this study is available from the corresponding author on reasonable request.

## Ethical statement

This study used de-identified, publicly available physiological databases and collected no new human or animal data.

## Author contributions

Khanh Duy Tran: Conceptualization; Methodology; Software; Validation; Formal analysis; Investigation; Data curation; Visualization; Writing—original draft; Writing—review & editing.

## Conflict of lnterest

The author declares no known competing financial interests or personal relationships that could have appeared to influence the work reported in this paper.

